# Association between disease severity and co-detection of respiratory pathogens in infants with RSV infection

**DOI:** 10.1101/2023.02.12.23285726

**Authors:** Gu-Lung Lin, Simon B Drysdale, Matthew D Snape, Daniel O’Connor, Anthony Brown, George MacIntyre-Cockett, Esther Mellado-Gomez, Mariateresa de Cesare, M Azim Ansari, David Bonsall, James E Bray, Keith A Jolley, Rory Bowden, Jeroen Aerssens, Louis Bont, Peter J M Openshaw, Federico Martinon-Torres, Harish Nair, Tanya Golubchik, Andrew J Pollard, the RESCEU Investigators

**Author notes:** Correspondence: G.-L. Lin, MD, DPhil, Oxford Vaccine Group, Centre for Clinical Vaccinology and Tropical Medicine, Churchill Hospital, Oxford OX3 7LE, UK. T. G. and A. J. P. contributed equally to this work.

## Abstract

**BACKGROUND:** Respiratory syncytial virus (RSV) is the leading cause of hospitalisation associated with acute respiratory infection in infants and young children, with substantial disease burden globally. The impact of additional respiratory pathogens on RSV disease severity is not completely understood.

**OBJECTIVES:** The objective of this study was to explore the associations between RSV disease severity and the presence of other respiratory pathogens.

**METHODS:** Nasopharyngeal swabs were prospectively collected from two infant cohorts: a prospective longitudinal birth cohort study and an infant cross-sectional study recruiting infants <1 year of age with RSV infection in Spain, the UK, and the Netherlands during 2017–20 [part of the REspiratory Syncytial virus Consortium in EUrope (RESCEU) project]. The samples were sequenced using targeted metagenomic sequencing with a probe set optimised for high-resolution capture of sequences of over 100 pathogens, including all common respiratory viruses and bacteria. Viral genomes and bacterial genetic sequences were reconstructed. Associations between clinical severity and presence of other pathogens were evaluated after adjusting for potential confounders, including age, gestational age, RSV viral load, and presence of comorbidities.

**RESULTS:** RSV was detected in 433 infants. Nearly one in four of the infants (24%) harboured at least one additional non-RSV respiratory virus, with human rhinovirus being the most frequently detected (15% of the infants), followed by seasonal coronaviruses (4%). In this cohort, RSV-infected infants harbouring any other virus tended to be older (median age: 4.3 vs. 3.7 months) and were more likely to require intensive care and mechanical ventilation than those who did not. *Moraxella, Streptococcus*, and *Haemophilus* species were the most frequently identified target bacteria, together found in 392 (91%) of the 433 infants (*S. pneumoniae* in 51% of the infants and *H. influenzae* in 38%). The strongest contributors to severity of presentation were younger age and the co-detection of *Haemophilus* species alongside RSV. Across all age groups in both cohorts, detection of *Haemophilus* species was associated with higher overall severity, as captured by ReSVinet scores, and specifically with increased rates of hospitalisation and respiratory distress. In contrast, presence of *Moraxella* species was associated with lower ReSVinet scores and reduced need for intensive care and mechanical ventilation. Infants with and without *Streptococcus* species (or *S. pneumoniae* in particular) had similar clinical outcomes. No specific RSV strain was associated with co-detection of other pathogens.

**CONCLUSION:** Our findings provide strong evidence for associations between RSV disease severity and the presence of additional respiratory viruses and bacteria. The associations, while not indicating causation, are of potential clinical relevance. Awareness of coexisting microorganisms could inform therapeutic and preventive measures to improve the management and outcome of RSV-infected infants.

## Introduction

Human respiratory syncytial virus (RSV) is the leading cause of hospitalisation associated with acute lower respiratory tract infection (LRTI) in infants and young children worldwide. The global RSV epidemic is estimated to cause 33 million LRTIs annually, leading to 3.6 million hospitalisations and around 100,000 RSV-attributable overall deaths in children under 5 years old, with mortalities predominantly in low- and middle-income countries^1^.

The standard of care for RSV infection is supportive management. Palivizumab, a monoclonal antibody for RSV prevention, has been used in young children at high risk for severe RSV LRTI, but it requires monthly administrations and its use is limited to higher-income settings due to prohibitive cost^2^. Nirsevimab, another monoclonal antibody with an extended half-life, has recently been authorised for use to prevent severe RSV LRTI with a single injection in neonates and infants during their first RSV season in the United Kingdom (UK) (licence numbers: PLGB 17901/0370–0371) and European Union^3^.

Infants normally harbour commensal microorganisms in the upper respiratory tract regardless of respiratory symptoms, including bacteria with pathogenic potential such as *Streptococcus pneumoniae* and *Haemophilus influenzae*^4^, and viruses such as enteroviruses and coronaviruses^5-8^. Simultaneous detection of multiple viruses is frequent in both healthy children^8^ and children with respiratory infections^8-14^, and virus-virus interactions can be either synergistic or antagonistic. A recent study showed that co-infection with influenza A virus and RSV can form hybrid viral particles in vitro, which evade neutralising antibodies and broaden receptor tropism^15^. In contrast, human rhinovirus (HRV) has been shown to interfere with influenza virus, reducing the chance of co-detection and co-circulation of both viruses at the individual and population levels, respectively^16^. Throughout the first year of life, there are constant changes in the respiratory microbiome with increasing biodiversity^5,17^. Colonisation with *Moraxella, Haemophilus*, or *Streptococcus* spp. and pneumococcal disease have been shown to be associated with viral respiratory infections^5,18,19^.

Molecular testing and typing of respiratory viruses and common bacterial pathogens have been increasingly used in the clinical setting. However, despite the high prevalence of colonisation and co-infection, the impact of other potential pathogens on the presentation of RSV infection (e.g., disease severity) remains to be understood.

In this study we applied high-throughput RNA-based targeted metagenomic sequencing to simultaneously recover over 100 bacterial and viral pathogens from respiratory samples prospectively collected from RSV-infected infants enrolled in two European multicentre studies. We sought to explore the associations between RSV disease severity and the presence of additional respiratory pathogens, focusing on bacteria and viruses. A better understanding of microbial factors associated with RSV disease severity will help direct therapeutic and preventive measures and improve the management and outcome of infants with RSV infection.

## Methods

### Study design and clinical data collection

Nasopharyngeal swabs were prospectively collected from infants under 1 year of age with primary RSV infection from the community and hospitals in Spain, the UK, and the Netherlands during the 2017–20 RSV seasons. These infants were enrolled in two clinical studies of the REspiratory Syncytial virus Consortium in EUrope (RESCEU) project, a European multicentre project investigating epidemiology, immunology, and virology of RSV infection. The two studies have been previously described in detail^20,21^.

In the longitudinal birth cohort study (ClinicalTrials.gov identifiers: NCT03627572), healthy term neonates (born at ≥37 weeks’ gestation) were enrolled. Key exclusion criteria were a history of major congenital defects (e.g., congenital heart and/or lung disease, genetic, immunological and/or metabolic disorders), acute severe medical conditions (e.g., sepsis or asphyxia), or receipt of immunoglobulin, monoclonal antibodies, or an investigational vaccine or medication against RSV. When participants developed respiratory symptoms during the RSV seasons before their first birthday, a nasopharyngeal swab was taken within 3 days of symptom onset and tested for RSV using point-of-care qualitative molecular testing on the Alere™ i RSV assay (Abbott, Illinois, United States). Regardless of the results, the swabs were immersed in M4RT^®^ transport medium after collection, aliquoted, and frozen at –80 °C until use.

In the infant cross-sectional study (ClinicalTrials.gov identifiers: NCT03756766), RSV-infected infants under 1 year of age were enrolled from the community within 4 days of symptom onset and from hospitals within 2 days of admission during the RSV seasons. Infants who were previously healthy and who had pre-existing medical conditions were both eligible for inclusion in this study. Key exclusion criteria were a history of RSV infection, receipt of immunoglobulin or monoclonal antibodies, or exposure to an RSV investigational vaccine or medication. RSV infection was diagnosed using the Alere™ i RSV assay in community settings or by routine antigen or polymerase chain reaction (PCR) tests at the hospital laboratory in hospital settings. A nasopharyngeal swab was collected at the time of enrolment for every participant. For hospitalised participants, daily nasopharyngeal swabs were also collected until hospital discharge. These swabs were also frozen at – 80 °C until use.

Demographic and clinical information was gathered in both studies. Clinical outcomes were evaluated by the ReSVinet score^22^, presence of fever, and requirement for hospitalisation, intensive care (i.e., admission to a high dependency unit or an intensive care unit), respiratory support, or invasive mechanical ventilation. The ReSVinet score accounts for seven clinical parameters, including feeding intolerance, medical intervention, respiratory difficulty, respiratory frequency, apnoea, general condition, and fever. The scores range from 0 to 20, with higher scores representing a more severe disease. Fever was defined as at least one episode of a tympanic or rectal temperature of ≥38 °C during the acute infection.

The studies were conducted in accordance with the provisions of the Declaration of Helsinki and were approved by the relevant authorities and ethics committees at each site: Hospital Clínico Universitario de Santiago de Compostela, Comité de Ética de la Investigación de Santiago-Lugo (no. 2017/395) in Spain; the University of Oxford, the Health Research Authority (no. 231136), the NHS National Research Ethics Service Oxfordshire Research Ethics Committee A (no. 15/SC/0335) and South Central – Hampshire A (no. 17/SC/0522) in the UK; and the Medical Ethical Committee, University Medical Center Utrecht (no. 17/563) in the Netherlands. The parents or guardians of all participants provided written, informed consent.

### Nucleic acid isolation and targeted metagenomic sequencing

Nucleic acid isolation and sequencing were performed as described previously^23-25^. The NucliSENS^®^ easyMAG^®^ system (BioMérieux, Marcy-l’Étoile, France) was used for automatic total nucleic acid extraction from 500 µL of each sample, following the manufacturer’s instructions. Sequencing libraries were constructed using the SMARTer^®^ Stranded Total RNA-Seq Kit v2 - Pico Input Mammalian (Takara Bio USA, California, United States), following a modified veSEQ-HIV protocol^23,24,26^. A 10-µL aliquot of each library was pooled together, and 750 ng of the pooled library was pulled down with a predesigned SureSelect RNA Target Enrichment multi-pathogen probe set (Agilent, California, United States). This probe set, *Castanet*, consisting of 120-mer oligonucleotides, was designed using the algorithm devised in the veSEQ protocol^27^. It targeted more than 100 potentially pathogenic bacteria and viruses, including both RSV-A and RSV-B (Table S1)^28^. The post-capture libraries were amplified with 16 cycles of PCR, and then purified using AMPure XP.

Sequencing was performed on the Illumina MiSeq platform (Illumina, California, US) or the Illumina NovaSeq 6000 system, generating paired-end reads. The MiSeq platform was used for a batch of ≤96 samples, and the NovaSeq 6000 system was used for a batch of 384 samples. Each 96-well sequencing plate also included one RSV-negative sample (collected from participants with acute respiratory symptoms, testing negative by the Alere™ i RSV assay) and one pure M4RT^®^ transport medium as negative controls. These negative controls were processed together with RSV-positive samples.

### Viral load measurement

Viral load was determined using reverse transcription quantitative PCR (RT-qPCR) assays performed at GSK as previously described^29^. The primers of this duplex RT-qPCR assay targeted the N gene for both RSV-A and RSV-B. The limit of detection was 304 and 475 copies/mL for RSV-A and RSV-B, respectively.

### Viral genome assembly and phylogenetic reconstruction

RSV genomes were reconstructed using shiver^30^ as previously described^23,24^. Briefly, reads were trimmed to remove adapters, random primers, and low-quality bases using Trimmomatic (v0.39)^31^ (option: Adapters:2:10:7:1:true LEADING:20 TRAILING:20 SLIDINGWINDOW:4:20 MINLEN:50). Trimmed reads were assembled into contigs using IVA (v1.0.8)^32^ and metaSPAdes (v3.14.1)^33^, and mapped to genotype-specific RSV references using shiver, with Bowtie 2 (v2.4.1)^34^ as the mapper (option: --very-sensitive-local --maxins 2000 --no-discordant --no-unal). Properly paired reads were retained and duplicates removed using Picard MarkDuplicates (v2.18.14, https://broadinstitute.github.io/picard/). Consensus sequences were generated by shiver, where base calling was supported by a minimum of two unique reads per position. RSV consensus sequences covering at least 70% of the coding sequences were used to reconstruct phylogenetic trees. For infants with multiple samples collected, only the sequence with the highest coverage was included. To put the study strains in a global context, we also included a subset of contemporaneous global RSV strains downloaded from GenBank on 4^th^ December 2020 with at least 70% coverage of the coding sequences, collected between 2015 and 2019. Genomic sequences were aligned using mafft (v7.490)^35^ with the FFT-NS-i method^36^. RAxML (v8.2.12)^37^ was used to reconstruct the maximum-likelihood phylogenies with the general time reversible nucleotide substitution model and gamma-distributed rate heterogeneity among sites. The R package ggtree (v2.2.4)^38^ was used for tree visualisation. Patristic distances, used to measure phylogenetic distances between sample pairs, were computed using the cophenetic function in the R package stats (v4.0.2). Maximum-likelihood phylogenies of enterovirus and human herpesvirus 6 (HHV-6) were reconstructed using RAxML with the same models as above.

### Respiratory viral genome reconstruction

The *Castanet* probe set^28^ is designed to cover the known genetic diversity of respiratory viruses, as well as phylogenetically informative sequences of respiratory-associated bacteria. It includes full genomes of viruses <40 kb in length, and 20 kb of sequence of longer genomes such as human herpesviruses (HHV), representing around 10% of the genome from the U23 to U37 genes. Although designed prior to the emergence of severe acute respiratory syndrome coronavirus 2 (SARS-CoV-2), this probe set includes sufficient coronavirus sequences to capture 23 kb of the 29-kb genome of SARS-CoV-2 (Fig. S1)^39^. The *Castanet* pipeline (Golubchik, https://github.com/tgolubch/castanet) was used to determine coverage of all target viruses.^28^ Reads originating from human cells were removed using BBMap (Bushnell, https://sourceforge.net/projects/bbmap/). The remaining host-depleted reads were mapped using Bowtie 2 to the references that covered known sequence variability of the target viruses. Read depth, genome coverage, and consensus sequences were generated for each virus from binary alignment/map (BAM) files.

Based on comparison with negative controls, viruses with more than 20–50 uniquely mapped reads and ≥30% genome recovery were considered present, and those with <10 unique reads were considered absent. Viruses fulfilling neither criterion were considered equivocal. Further criteria varied between viruses due to different efficiencies of the target enrichment and sequence specificities; detailed criteria for each virus are shown in Table S2.

Enterovirus genotyping was performed using the web-based Enterovirus Typing Tool^40^ (https://www.rivm.nl/mpf/typingtool/enterovirus/), based on phylogenetic analysis of the VP1 gene. The genotypes of HHV-6 were determined by phylogenetic analysis of the HHV-6 partial genome consensus sequences and reference strains. These references included HHV-6A (strains: U1102, GS, and AJ; accession numbers: NC_001664, KC465951, and KP257584), HHV-6B (strains: Z29 and HST; accession numbers: NC_000898 and AB021506), chromosomally integrated HHV-6A (accession numbers: MG894371 and KY315540), and chromosomally integrated HHV-6B (accession numbers: KY316051 and MG894376).

### Respiratory bacterial reconstruction

The *Castanet* probe set also covered the known variation of 53 *rps* genes of the target bacteria. These genes encode the bacterial ribosome protein subunits, and their allele characterisation can be used to classify bacteria into groups at all taxonomic and most typing levels^41^. A database containing catalogued *rps* gene variation has been developed for ribosomal multilocus sequence typing (rMLST)^41^.

Reconstruction of the bacterial genetic sequences was based on the rMLST scheme. Contig sequences assembled by metaSPAdes were matched against all known alleles for each *rps* gene to find exact matches. The PubMLST multi-species isolate database, integrating curated allelic and species information, was then searched to identify the matching species (algorithm available at https://pubmlst.org/species-id)^41^. The PubMLST RESTful application programming interface was applied to efficiently analyse all samples^42,43^.

Bacteria supported by at least two exact and unique allelic matches were considered present. A ‘unique’ allelic match is an allelic pattern that is linked to exactly one taxon at its lowest taxonomic rank or two bacteria with one being an unspecified species of the same genus as the other specified species (e.g., if *Moraxella* sp. and *Moraxella catarrhalis* were both linked to an allelic pattern, *Moraxella catarrhalis* was considered a unique match). Bacteria supported by only one exact and unique match were considered equivocal. The rest were considered absent. The number of reads assigned to a certain species was determined by taxonomic classification of the trimmed reads using Kraken 2 (v0.39)^44^. A Krona chart was used to visualise the composition of identified bacteria.^45^

### Statistical analyses

For infants with multiple samples collected, species that were detected in at least one sample collected while RSV was detectable or within 7 days of enrolment (whichever is longer) were considered present; species that were not detected in any sample collected while RSV was detectable or within 7 days of enrolment (whichever is longer) were considered absent; the rest were considered equivocal. Infants with an equivocal presence of a species were excluded from clinical outcome comparisons.

Phylogenetic signal for a co-detected species on RSV phylogenies was evaluated using the *D* statistic based on a permutation testing framework^46^. An equivocal presence status was considered absence in this analysis. A *D* value of 0 indicates a strong phylogenetic signal under the Brownian motion model of evolution (i.e., conserved trait evolution), and a value of 1 indicates no phylogenetic signal (i.e., a random phylogenetic distribution). A negative *D* value indicates that the binary trait is more conserved than the expectations of the Brownian motion model, whereas a value greater than 1 indicates phylogenetic overdispersion. The *D* statistical tests were performed using the phylo.d function in the R package caper (v1.0.1).

When comparing clinical outcomes between different groups of infants, multiple linear regression (for continuous outcome variables), multivariable logistic regression (for dichotomous outcome variables), and proportional odds ordered logistic regression (for ordered outcome variables) were used to adjust for covariates. Covariates included age, gestational age, sex, comorbidity, sampling season and country, study, viral subgroup, peak RSV read count, and the duration between symptom onset and sampling. Models with different combinations of covariates were tested, and the model with the lowest Akaike information criterion (AIC) was selected. A post hoc adjustment for multiple comparisons with the Benjamini–Hochberg method was applied to determine false discovery rate– corrected Q values in all clinical outcome comparisons. The effect size of the presence of a pathogen on a clinical outcome was evaluated using Cohen’s f^2^. An f^2^ value between 0.02 and <0.15 represents a small effect size; a value between 0.15 and <0.35, medium; and a value of ≥0.35, large^47^. All statistical analyses were performed using R (v4.0.2)^48^.

## Results

### Study and sample populations

Overall, 839 nasopharyngeal swabs from a total of 440 RSV-infected infants, enrolled from Spain, the UK, and the Netherlands during 2017–2020, were sequenced (Table S3). One infant’s sample could not be sequenced, and no RSV reads were recovered from six infants’ samples (n = 7), five of which had undetectable viral load. These seven infants and eight samples were excluded from further analysis. A total of 125 hospitalised infants in the infant cross-sectional study had more than one swab collected (mean ± SD, 4.2 ± 2.5).

The median age of the remaining 433 infants was 4.1 months [interquartile range (IQR), 1.9–7.5 months] at the time of the RSV infection. Females accounted for 45%. Nine percent of the infants had pre-existing medical conditions, including prematurity with or without bronchopulmonary dysplasia, ventricular septal defect, and other congenital abnormalities. Younger infants tended to have a higher ReSVinet score and were more likely to require hospitalisation, intensive care, respiratory support, and mechanical ventilation than older infants, whereas a greater proportion of older infants had a fever than younger infants (<3 months vs. 3 to <6 months vs. 6 to <12 months) (Table S4). Due to the differences in study design, infants in the infant cross-sectional study were likely to be younger and have more severe disease than those in the longitudinal birth cohort study (Table S5).

We previously showed^23^ that the number of uniquely mapped RSV reads from our protocol is highly correlated with RSV viral load (Pearson correlation, R^2^ = 0.78, P = 5×10^−197^; Fig. S2). Thus, we used the number of unique RSV reads as a surrogate for viral load for the analyses that follow. Of the 831 samples where RSV sequencing was possible, all 597 (72%) samples with at least 70% of the RSV genome reconstructed had a median of 11,851 unique RSV reads (range, 285 to 810,113), corresponding to a median viral load of 1.2×10^7^ copies/mL (range, 505 to 4.3×10^9^).

Similar proportions of the 433 infants had RSV-A and RSV-B (220 vs. 204; the remaining 9 could not be genotyped due to evidence of mixed infection or contamination and were excluded from genomic analyses). RSV-B dominated the 2017–18 and 2018–19 RSV seasons, accounting for 71% and 55% of the infections, respectively, whereas RSV-A was the predominant subgroup during the 2019–20 RSV season, accounting for 66% of the infections (Table S6). Phylogenetic analyses classified all RSV-A strains into genotype ON1 and all RSV-B strains into genotype BA (Fig. S3). There was no evidence to support differences in severity between RSV-A and RSV-B infections (Table S7).

### RSV and co-detected respiratory viruses

Co-infection with multiple respiratory viruses, defined as co-detected presence of at least two respiratory viruses, was common among the infants in this study. At least one respiratory virus in addition to RSV was recovered in 106 (24%) of the 433 RSV-infected infants, including 10 infants with two additional viruses (Fig. 1). Non-RSV viruses were present at substantial viral load: in the quantitative sequencing method we used, over 90% of the non-RSV viruses had sufficient unique reads to enable reconstruction of at least 50% of their genome, consistent with viral loads in a range similar to that of RSV and suggestive of active replication/infection (Fig. S4). RSV-infected infants with any viral co-infection tended to be older than those without [median (IQR), 4.3 (2.3–8.2) vs. 3.7 (1.7–6.9) months; Mann–Whitney U test, P = 0.031] (Table S8 and Fig. S5). There was no correlation between the viral load of non-RSV viruses and RSV viral load or genotype, and likewise no correlation between viral load of non-RSV viruses and infant age, sex, or clinical outcome.

**Figure 1.**
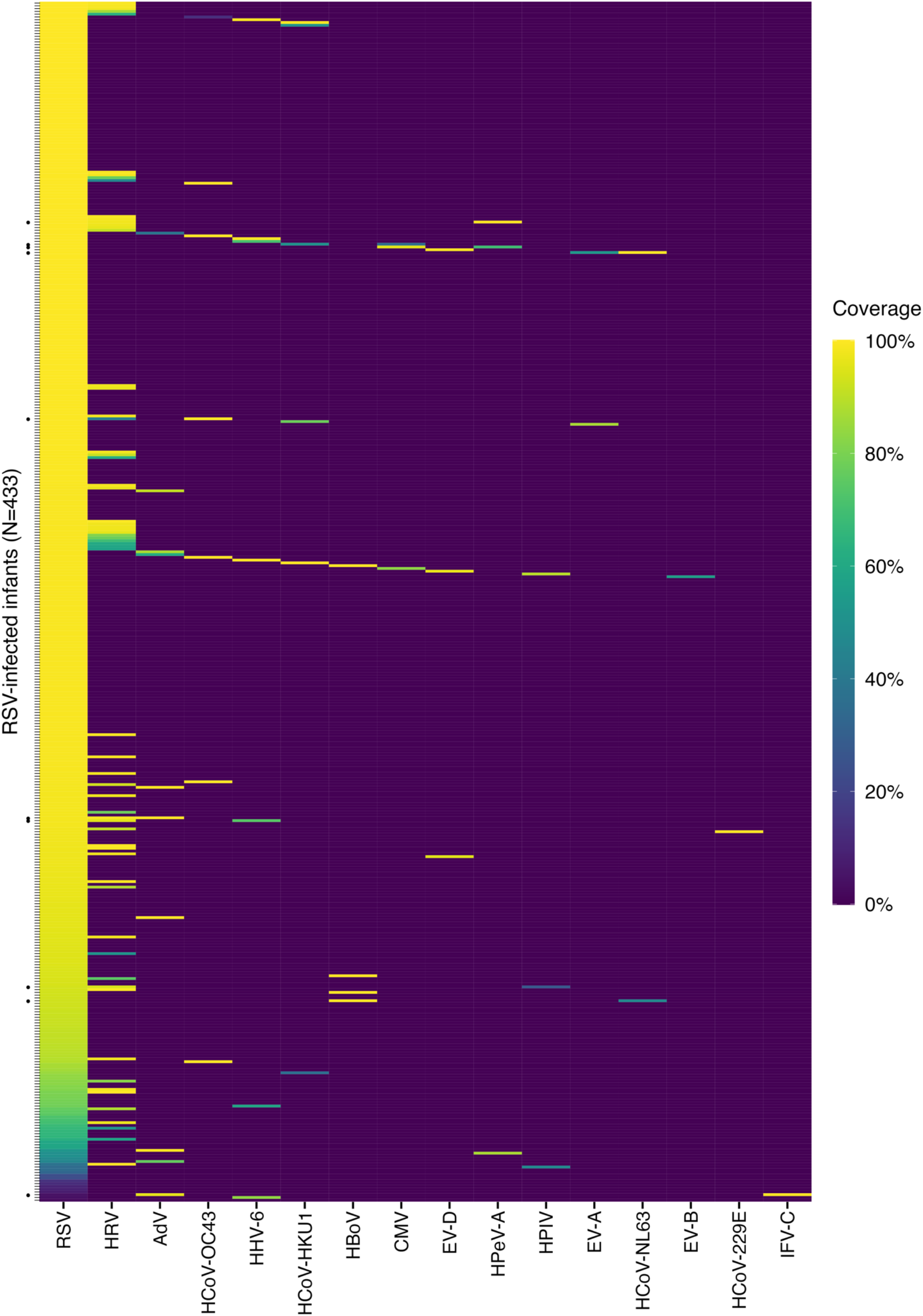
Genome coverage of all detected viruses in RSV-infected infants. Each row represents an individual infant. Dots on the Y axis are the 10 infants who had two other viruses identified in addition to RSV. Genome coverage is illustrated by the coloured gradient, defined as proportion of targeted genome that was reconstructed. Viruses are ordered in decreasing prevalence from left to right. RSV denotes respiratory syncytial virus; HRV, human rhinovirus; AdV, adenovirus; HCoV, human coronavirus; HBoV, human bocavirus; CMV, human cytomegalovirus; EV, enterovirus; HPeV-A, human parechovirus A; HPIV, human parainfluenza virus; and IFV-C, influenza C virus.

Human rhinovirus (HRV) was the virus most frequently found alongside RSV (63/433 infants, 15%), followed by seasonal human coronaviruses (HCoVs, 4%) and adenoviruses (2%) (Table S9). We did not find any SARS-CoV-2 in 76 swabs collected from 35 infants in January and February 2020. VP1 genotyping of the HRV detected in the 63 infants showed that HRV-A, HRV-B, and HRV-C accounted for 41% (26/63), 29% (18/63), and 30% (19/63), respectively, as well as three cases of enterovirus D68 (Fig. S6). Human herpesvirus 6 (HHV-6) was detected in seven (2%) infants, including two HHV-6A and five HHV-6B (Fig. S7). Phylogenetic segregation of HHV-6A was strongly suggestive of chromosomally integrated HHV-6A in our samples^49^.

We examined the impact of additional viruses on markers of disease severity after adjusting for covariates, including age, gestational age, RSV viral load, and presence of comorbidities (Fig. 2). There was strong evidence that infants with viral co-infection were more likely to require intensive care and mechanical ventilation than those without (Fig. 2, Table S8). Analysing each non-RSV virus family separately (i.e., enterovirus, HRV, HCoV, adenovirus, and HHV-6) showed that infants co-infected with HCoV tended to be older than those without [median (IQR), 7.7 (3.84–10.0) vs. 4.0 (1.8– 7.4) months; Mann–Whitney U test, P = 0.012]. There was also some evidence that infants co-infected with HHV-6 were more likely to require intensive care than those without (OR, 10.4; 95% CI, 1.7– 86.9; P = 0.015; Cohen’s f^2^ = 0.184), but limited sample numbers of HHV-6 preclude a robust comparison (not significant after adjusting for multiple comparisons).

**Figure 2.**
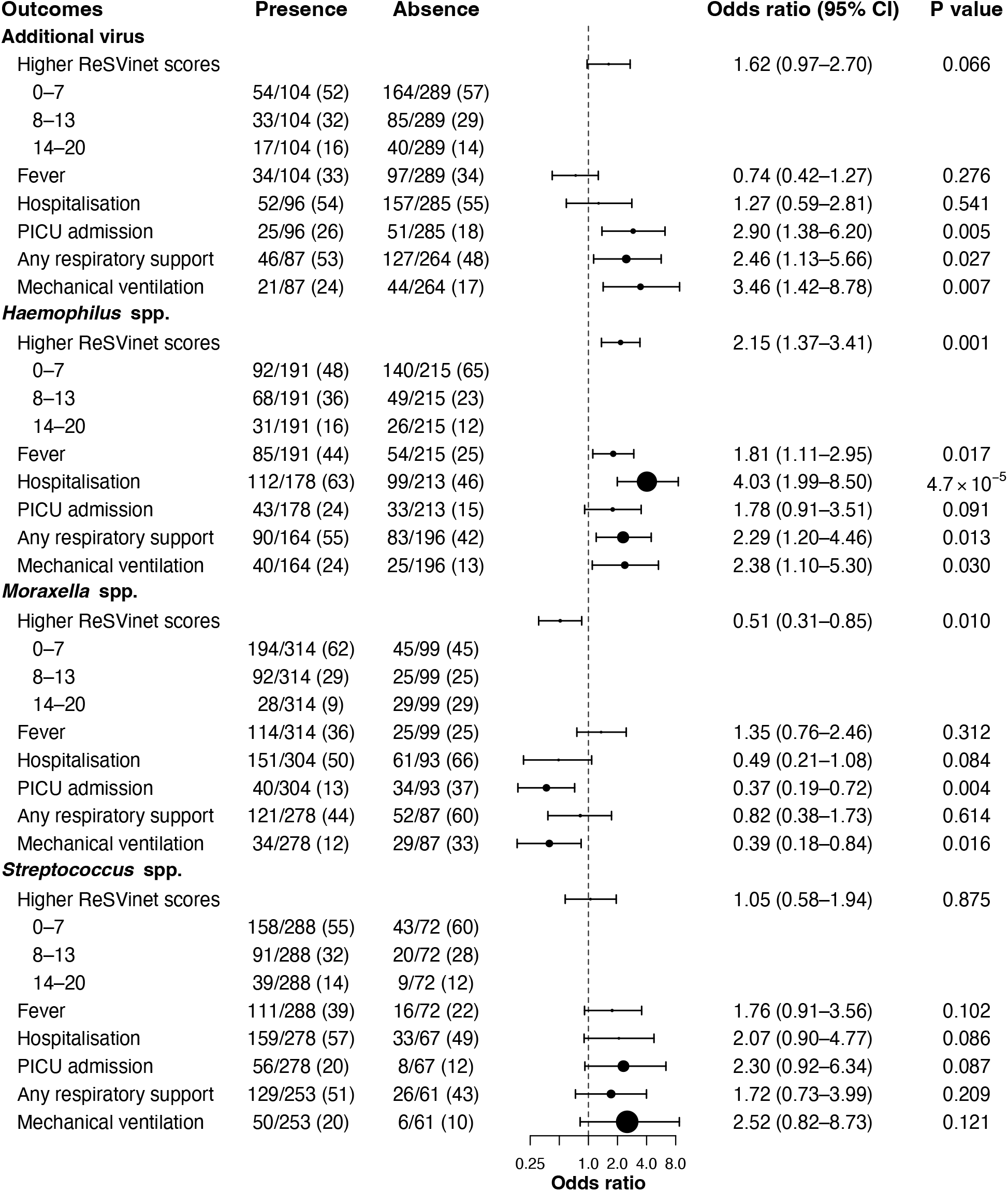
Clinical outcomes by presence and absence of pathogen groups. Odds ratios were adjusted for covariates including age, gestational age, sex, comorbidity, sampling season and country, study, RSV subgroup, peak RSV read count, and the duration between symptom onset and sampling using ordered logistic regression or multivariable logistic regression. Data in presence and absence are shown as no./total no. (%). Black dots represent the mean odds ratio, sized in proportion to Cohen’s f^2^, ranging from 0.0009 (fever for *Streptococcus* spp.) to 0.0958 (mechanical ventilation for *Streptococcus* spp.). CI denotes confidence interval, and PICU paediatric intensive care unit.

Figure 3 shows the distribution of the non-RSV viruses on the RSV phylogenies. There was no significant phylogenetic signal for the presence of these viruses on the RSV phylogenies with *D* values ranging from 0.80 to 1.33, indicating no specific RSV clade was linked with the presence of other viruses (Table S10).

**Figure 3.**
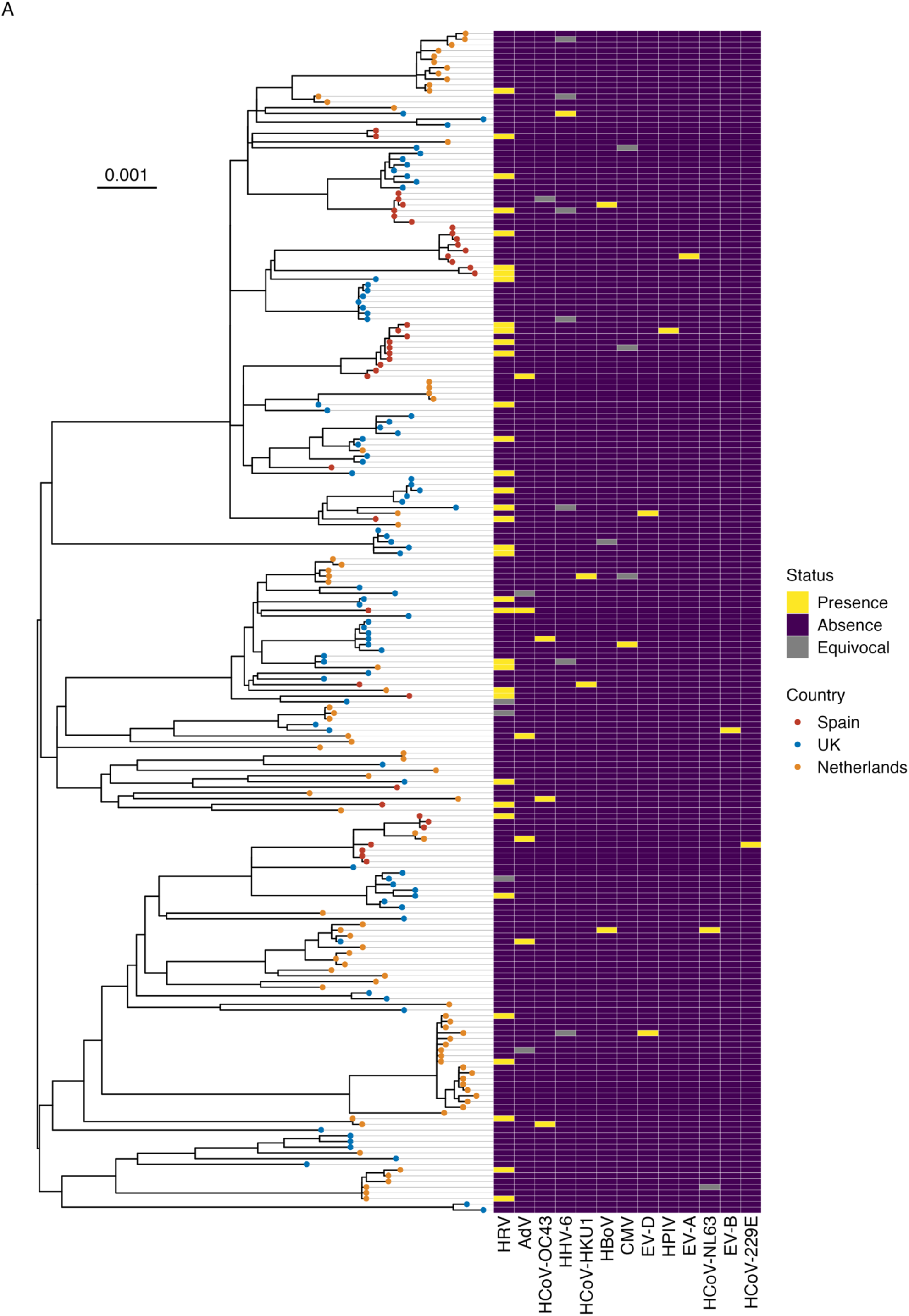
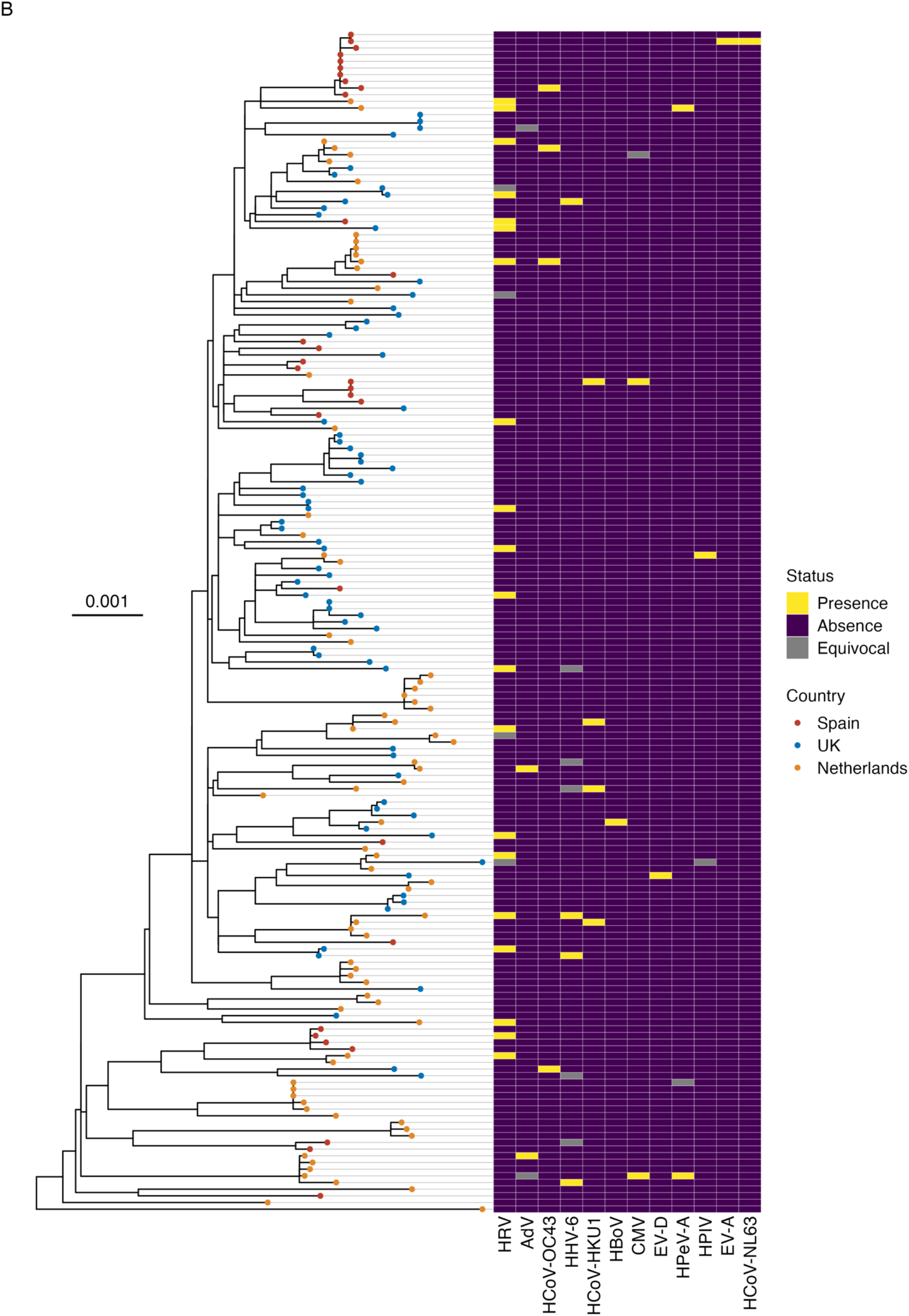
Distribution of co-detected viruses on the (A) RSV-A and (B) RSV-B maximum-likelihood phylogenies. The phylogenies were reconstructed from samples in which at least 70% of the RSV coding sequences were recovered, using RAxML with the general time reversible nucleotide substitution model and gamma-distributed rate heterogeneity among sites. For infants with multiple samples collected, only the sample with the highest coverage was included. The trees were midpoint rooted. The sampling country of each strain is illustrated by tip colour. The scale bars represent the number of nucleotide substitutions per site. Human parechovirus A (HPeV-A) was not detected with any of the RSV-A strains; enterovirus B (EV-B) and human coronavirus 229E (HCoV-229E) were not detected with any of the RSV-B strains. HRV denotes human rhinovirus; AdV, adenovirus; HHV-6, human herpesvirus 6; HBoV, human bocavirus; CMV, human cytomegalovirus; and HPIV, human parainfluenza virus.

### RSV and co-detected bacterial pathogens

We detected target bacterial species in the vast majority of infants (406/433, 94%). Among the 21/27 infants with no detected target bacteria for whom information on antibiotic usage was available, 10 did not receive any antibiotics during the RSV infection, 10 had received antibiotics before sample collection, and one received antibiotics without information on the timing of antibiotic dosing. These 27 infants had a mean ± SD peak RSV read count of 3.7 ± 1.2 log_10,_ falling in the lower quartile of the study population (Fig. S4), suggesting these may have been poorer quality swabs.

Excluding these 27 infants, the mean ± SD number of target bacterial species was 3.4 ± 2.0, with *Moraxella* (76% of all 433 RSV-infected infants), *Streptococcus* (68%), and *Haemophilus* (45%) being the most frequently found genera (Table S11, Figs. S8 and S9). These three genera together were found in 392/433 infants (91%). Among each of these genera, *M. catarrhalis, S. pneumoniae*, and *H. influenzae* were the most commonly identified species, and had read numbers consistent with substantial bacterial load (Fig. S4). Infants with any of these three bacterial genera were older than those without [median (IQR), 4.6 (1.9–7.7) months vs. 2.0 (1.1–3.4) months; Mann–Whitney U test, P = 0.002].

Five of the recovered bacterial species were further classified down to the ribosomal sequence type (rST) level in some samples: *M. catarrhalis, S. pneumoniae, H. influenzae, Escherichia coli*, and *Neisseria meningitidis*. Most rSTs were unique in the dataset, consistent with extensive bacterial genetic diversity at the population level. Only a few isolates shared the same rST—*M. catarrhalis* rST 105842 was found in two infants; each of the *S. pneumoniae* rSTs 644, 12187, 13906, and 104663 were found in two infants; and each of the *H. influenzae* rSTs 24162, 49634, 89110, and 133371 were also found in two infants (Table S12).

Similarly to the analysis of viral co-infections, we investigated the correlation between disease severity and the presence of *Moraxella, Haemophilus*, and *Streptococcus*, after adjusting for confounders as above. Compared with infants without any *Haemophilus* spp., infants with *Haemophilus* spp. were more likely to have a higher ReSVinet score and fever and require hospitalisation, respiratory support, and mechanical ventilation (Figs. 2 and 4A, Table S13). Although the effect size of *Haemophilus* presence on most of these clinical outcomes was small (Cohen’s f^2^ between 0.02 and <0.15) (Table S13), presence of *Haemophilus* spp. correlated with around a 2.6-point increase in ReSVinet scores across all age groups, in both cohorts that made up the study population (Fig. 4A). As expected, ReSVinet scores correlated with age for the infants enrolled in the infant cross-sectional study (Pearson correlation, P = 3.3×10^−9^) but not for those in the longitudinal birth cohort study (P = 0.44) (Fig. 4A). This was because infants in the infant cross-sectional study were recruited on presentation with an RSV infection, while those in the longitudinal birth cohort study were prospectively followed up to detect RSV positivity and thus more likely to include mild cases.

**Figure 4.**
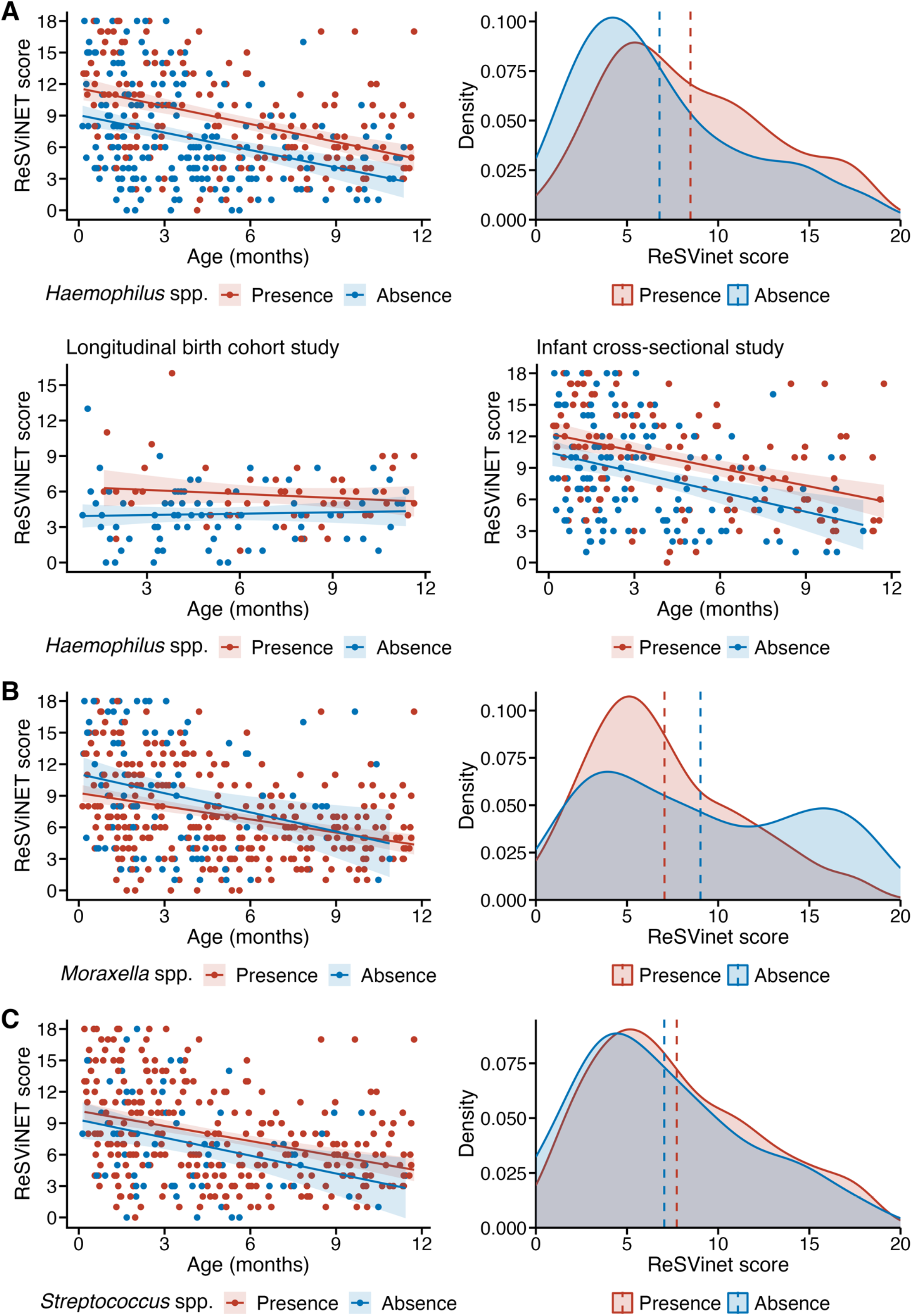
Relationships between ReSVinet score and age, and density plots of ReSVinet score, stratified by presence status of (A) *Haemophilus* spp., (B) *Moraxella* spp., and (C) *Streptococcus* spp. Results from the longitudinal birth cohort study and infant cross-sectional study are also shown separately in panel A. On the left panel, each dot represents an individual infant, and the lines are the regression lines with the shaded area representing the 95% confidence interval. On the right panel, dashed lines represent the mean ReSVinet score.

In contrast to the observation of higher ReSVinet scores with *Haemophilus* spp., infants with *Moraxella* spp. tended to have a lower ReSVinet score (especially in infants under 6 months of age; Fig. 4B) and were less likely to require intensive care and mechanical ventilation than those without (Fig. 2). Infants with and without *Streptococcus* spp., or *S. pneumoniae* in particular, had similar clinical outcomes (Figs. 2 and 4C).

Given the different direction of effect of *Haemophilus* spp. vs. *Moraxella* spp. on disease severity, we hypothesised that antibiotic administration may have been higher among infants with *Haemophilus* spp., potentially indicating secondary bacterial infection. Information on the use of antibiotics was available in 361 RSV-infected infants, of which 25% received antibiotics during their RSV episode. Consistent with our hypothesis, a higher proportion of infants with *Haemophilus* spp. received antibiotics than those without (33% vs. 20%; Chi-square test, P = 0.010); and of these receiving antibiotics, 46% had received antibiotics before the first sample where *Haemophilus* spp. were identified. In contrast, a lower proportion of infants with *Moraxella* spp. received antibiotics than those without (19% vs. 43%; P = 2.2×10^−5^). The rates of receiving antibiotics were similar in infants with and without *Streptococcus* spp. (22% vs. 29%; P = 0.363), but fewer infants with *S. pneumoniae* received antibiotics than those without (19% vs. 30%; P = 0.019).

Figure S10 shows the distribution of commonly co-detected bacterial species (namely *M. catarrhalis, S. pneumoniae, H. influenzae*, and *Staphylococcus aureus*) on the RSV phylogenies. *S. pneumoniae* was non-randomly distributed across the RSV-B phylogeny (P = 0.026) with a *D* value of 0.77, but the distribution was significantly different from the expectations under Brownian motion (P <0.001), indicating a low phylogenetic signal (Table S10). There was no significant phylogenetic signal between the RSV phylogenies and the presence of other commonly co-detected bacteria, with *D* values ranging from 0.76 to 1.01.

## Discussion

Using RNA-based quantitative sequencing, we captured a broad range of respiratory pathogens in infants with RSV, with the aim of detecting additional viral and bacterial pathogens and examining correlates of disease severity. We found strong genomic evidence for presence of at least one additional virus in a high proportion (24%) of the 433 infants with active RSV infection, in keeping with previous reports of viral co-infection rates of 15–34% in children of all ages with respiratory infections^50-53^. This is likely a conservative estimate of the true co-infection rate, as our stringent detection criteria would necessarily miss very low-viral load co-infections. We additionally assessed the presence of respiratory bacteria with high pathogenic potential, and found that the three most commonly detected genera—*Moraxella, Streptococcus*, and *Haemophilus*—were detected in an overwhelming majority (94%) of the tested samples.

In our study population, presence of respiratory viruses in addition to RSV correlated with increased need for intensive care and mechanical ventilation, but the overall impact of viral co-infection on severity was small. Likewise, the presence of most bacterial species had no negative effect on severity. Indeed, presence of *Moraxella*, the most commonly detected bacteria, was associated with less severe presentation, with reduced probability of requiring intensive care and mechanical ventilation. In contrast, there was strong evidence that the presence of *Haemophilus* spp. was associated with severe RSV disease independent of age, study cohort, or RSV genotype, and this effect was demonstrated by multiple clinical outcome variables.

There is a general perception that an inverse relationship exists between age and rates of viral co-infections—infants and children have a higher rate of viral co-infections than adults^50,53-55^. However, at the lower extreme of age, under 12 months, we observed that older infants were more likely to have multiple respiratory viruses than younger infants. This may be due to presence of maternal antibodies in the youngest infants and a higher probability of social contacts (e.g., attending nursery) in older infants, exposing them to more respiratory viruses.

In our dataset, HRV was the most commonly co-infecting virus, found in 15% of the RSV-infected infants. The Pneumonia Etiology Research for Child Health (PERCH) study showed that HRV was the second most common virus causing severe pneumonia in children after RSV, and the most common virus in healthy children or those without severe pneumonia^8^. Although we did not look at HRV in the general population, it has been shown that the presence of RSV reduces the chance of concurrent HRV infection^56,57^, and the rate of HRV detection in our study was in accordance with that in previous studies which demonstrated interference between RSV and HRV^56^. Other viruses were co-detected with RSV in a further 9% of the infants. These viruses were taxonomically diverse and no single virus apart from HRV was found in >5% of the infants.

Co-detected viruses could represent either active replication of an additional virus (either newly acquired or reactivated) or leftover nucleic acid from a previous infection; the additional virus may be either as a non-pathogenic ‘bystander’ or a true disease-causing agent. Although not definite, unique read count as a surrogate for viral load may provide a clue to differentiate between these possibilities. Co-detected viruses with low viral load may come from prolonged shedding from a previous infection, whereas those with high viral load are more likely to cause symptoms. In this study, we focused on viruses with evidence of substantial viral load based on read numbers, as being consistent with active replication and therefore possible co-infection.

Among the bacterial genera we tested, the presence of *Haemophilus* spp. was significantly associated with severe RSV disease, consistent with the results of previous studies^5,58,59^. A recent study also related high levels of *Haemophilus* spp. with early recurrence of respiratory symptoms following a lower respiratory tract infection^60^. In contrast, the presence of *Streptococcus* spp. (or *S. pneumoniae* specifically) was not associated with any of the tested clinical outcomes in our study. Acquisition, but not carriage, of *S. pneumoniae* by infants has been associated with medically attended infectious illness^61^. Contrary to our study, a previous study showed an association between *Streptococcus* abundance and an increased likelihood of hospitalisation for RSV infection^58^. *Haemophilus* spp. may have caused secondary bacterial infection in the RSV-infected infants since a greater proportion had fever and received antibiotics than those without *Haemophilus* spp. In contrast, the evidence for clinically suspected secondary bacterial infection with *Streptococcus* spp. or *Moraxella* spp. was insufficient. Overrepresentation of *H. influenzae* and *Streptococcus* spp. in RSV infection has been associated with enhanced Toll-like receptor (TLR) signalling^58^, mucosal chemokine (C-X-C motif) ligand 8 (CXCL8, also known as IL-8) responses^62^, IL-17A signalling^58^, and inflammatory responses^63^. CXCL8 and IL-17A signalling contribute to macrophage and neutrophil activation and recruitment^58^, inducing bronchoalveolar neutrophil infiltration^64^.

Despite being the most frequently found bacteria, *Moraxella* spp. were associated with less severe RSV infection that did not require intensive care or mechanical ventilation in our study, suggesting a potential protective effect of these microorganisms on RSV infection or their role as a non-pathogenic ‘bystander.’ Previous studies also showed that *Moraxella* spp. were more frequently found in outpatients than hospitalised children^58^ and children without severe pneumonia (including healthy children) than those with^8^, and significantly associated with fever in infants with RSV lower respiratory tract infections^5^. Although fever is one of the parameters in the ReSVinet score, we demonstrated elsewhere that it did not correlate with the other clinical outcome measures in a subset of these RSV-infected infants^25^. In this study, older infants were more likely to have a fever but generally presented with a milder RSV infection than younger infants. Further studies are required to evaluate the underlying immune mechanisms contributing to the potential protective effect of *Moraxella* spp.

Our study has several limitations. Due to regional and age-specific differences in respiratory flora, results generated from this study may not be generalisable to infants in low- and middle-income countries or different age groups (e.g., older adults). Although our method covers a comprehensive list of respiratory-associated viruses, co-detection does not imply capacity to initiate infection in the absence of RSV; for instance, we cannot determine whether HHV-6 detected in the infants was a primary respiratory pathogen or reactivation of a latent infection. More broadly, the associations observed in this study between disease severity and co-detection of respiratory pathogens in RSV-infected infants should be interpreted with caution and do not indicate causation. Such an indication would only come from a therapeutic trial. In addition, the rMLST scheme included in our method was able to classify bacteria to the species level or even the rST level, but the link between rST and serotype is currently unknown, which makes it difficult to classify detected *S. pneumoniae* and *H. influenzae*. Lastly, although multiple commensal and pathogenic viruses and bacteria were recovered, we did not attempt to characterise the whole microbial community. Microbiome analyses using samples collected in the RESCEU studies are ongoing.

This study deepens our understanding of the associations between RSV disease severity and the co-detected respiratory pathogens. Targeted metagenomic sequencing with the *Castanet* probe set enables simultaneous detection of combinations of bacterial and viral pathogens that may be of particular concern in RSV and other infections. Our results can assist in identifying patients at risk for severe RSV infection, directing the development of therapeutic and preventive measures, and improving the management and outcomes of individuals infected with RSV. Future studies evaluating the transcriptomic and immunological profiles of RSV infection should also consider the presence of other bacterial and viral pathogens.

## Data Availability

All raw sequencing reads of the samples included in this study are accessible at the European Nucleotide Archive under the study accession PRJEB34042 (https://www.ebi.ac.uk/ena/data/view/PRJEB34042). The RSV consensus sequences included in the phylogenetic analyses have been deposited in GenBank. The accession numbers can be found in Table S14.

## RESCEU investigators

Harish Nair, Harry Campbell, Steve Cunningham (University of Edinburgh); Philippe Beutels (Universiteit Antwerpen); Louis Bont, Joanne Wildenbeest, Debby Bogaert (University Medical Centre Utrecht); Andrew Pollard, Matthew Snape, Simon Drysdale, Gu-Lung Lin, Daniel O’Connor, Elizabeth Clutterbuck, Joseph McGinley, Hannah Robinson (University of Oxford); Peter Openshaw, Ryan Thwaites, Dexter Wiseman (Imperial College London); Federico Martinon-Torres, Alberto Gómez-Carballa, Carmen Rodriguez-Tenreiro, Irene Rivero-Calle, Ana Dacosta-Urbieta, Sara Pischedda (Servicio Galego de Saude); Terho Heikkinen (University of Turku and Turku University Hospital); Adam Meijer (National Institute for Public Health and the Environment); Thea K Fischer (Statens Serum Institut); Maarten van den Berge (University of Groningen); Carlo Giaquinto (PENTA Foundation); Michael Abram (AstraZeneca); Kena Swanson (Pfizer); Bishoy Rizkalla (GSK); Charlotte Vernhes, Scott Gallichan (Sanofi Pasteur); Jeroen Aerssens, Deniz Öner (Janssen); Veena Kumar (Novavax); Eva Molero (Team-It Research).

## Acknowledgements

This work was supported by the National Institute for Health Research (NIHR) Oxford Biomedical Research Centre, the NIHR Thames Valley and South Midlands Clinical Research Network, the British Research Council, and the Respiratory Syncytial Virus Consortium in Europe (RESCEU) project. RESCEU has received funding from the Innovative Medicines Initiative 2 Joint Undertaking (grant number 116019). This Joint Undertaking receives support from the European Union Horizon 2020 Research and Innovation Program and European Federation of Pharmaceutical Industries and Associations.

## Author contributions

G.-L. L., T. G., and A. J. P. conceived and designed the work. G.-L. L, S. B. D., M. D. S., J. A., L. B., P. J. M. O., F. M.-T., H. N., and A. J. P. conducted and supervised the clinical studies. M. A. A. designed the probe set that was used for capture. J. E. B. and K. A. J. designed the algorithm for bacterial identification. M. d. C., D. B., and R. B. designed the sequencing protocol. G.-L. L., A. B., G. M.-C., E. M.-G., and M. d. C. performed the experiments. G.-L. L., T. G., D. O’C., M. A. A., J. E. B., and K. A. J. analysed and interpreted the data. G.-L. L. drafted the manuscript, and all other authors critically revised it. All authors have approved the submitted version and agreed to submit the manuscript. All authors agree to be accountable for all aspects of the work in ensuring that questions related to the accuracy and integrity of any part of the work are appropriately investigated and resolved.

## References

1. Li Y, Wang X, Blau DM, et al. Global, regional, and national disease burden estimates of acute lower respiratory infections due to respiratory syncytial virus in children younger than 5 years in 2019: a systematic analysis. Lancet 2022;399:2047–64.

2. American Academy of Pediatrics. Updated guidance for palivizumab prophylaxis among infants and young children at increased risk of hospitalization for respiratory syncytial virus infection. Pediatrics 2014;134:e620–38.

3. European Medicines Agency. Beyfortus. 2022. Accessed on 23 Jan, 2023, at https://www.ema.europa.eu/en/medicines/human/EPAR/beyfortus

4. Wouters I, Desmet S, Van Heirstraeten L, et al. How nasopharyngeal pneumococcal carriage evolved during and after a PCV13-to-PCV10 vaccination programme switch in Belgium, 2016 to 2018. Euro Surveill 2020;25:1900303.

5. Teo SM, Mok D, Pham K, et al. The infant nasopharyngeal microbiome impacts severity of lower respiratory infection and risk of asthma development. Cell Host Microbe 2015;17:704–15.

6. Bosch AA, Biesbroek G, Trzcinski K, Sanders EA, Bogaert D. Viral and bacterial interactions in the upper respiratory tract. PLoS Pathog 2013;9:e1003057.

7. Cebey-López M, Herberg J, Pardo-Seco J, et al. Viral co-infections in pediatric patients hospitalized with lower tract acute respiratory infections. PLoS One 2015;10:e0136526.

8. Pneumonia Etiology Research for Child Health (PERCH) Study Group. Causes of severe pneumonia requiring hospital admission in children without HIV infection from Africa and Asia: the PERCH multi-country case-control study. Lancet 2019;394:757–79.

9. Martin ET, Kuypers J, Wald A, Englund JA. Multiple versus single virus respiratory infections: viral load and clinical disease severity in hospitalized children. Influenza Other Respir Viruses 2012;6:71–7.

10. Aberle JH, Aberle SW, Pracher E, Hutter HP, Kundi M, Popow-Kraupp T. Single versus dual respiratory virus infections in hospitalized infants: impact on clinical course of disease and interferon-gamma response. Pediatr Infect Dis J 2005;24:605–10.

11. Yoshida LM, Suzuki M, Nguyen HA, et al. Respiratory syncytial virus: co-infection and paediatric lower respiratory tract infections. Eur Respir J 2013;42:461–9.

12. Mansbach JM, Piedra PA, Teach SJ, et al. Prospective multicenter study of viral etiology and hospital length of stay in children with severe bronchiolitis. Arch Pediatr Adolesc Med 2012;166:700–6.

13. Greer RM, McErlean P, Arden KE, et al. Do rhinoviruses reduce the probability of viral co-detection during acute respiratory tract infections? J Clin Virol 2009;45:10–5.

14. Mazur NI, Bont L, Cohen AL, et al. Severity of respiratory syncytial virus lower respiratory tract infection with viral coinfection in HIV-uninfected children. Clin Infect Dis 2017;64:443–50.

15. Haney J, Vijayakrishnan S, Streetley J, et al. Coinfection by influenza A virus and respiratory syncytial virus produces hybrid virus particles. Nat Microbiol 2022;7:1879–90.

16. Nickbakhsh S, Mair C, Matthews L, et al. Virus-virus interactions impact the population dynamics of influenza and the common cold. Proc Natl Acad Sci U S A 2019;116:27142–50.

17. Prevaes SM, de Steenhuijsen Piters WA, de Winter-de Groot KM, et al. Concordance between upper and lower airway microbiota in infants with cystic fibrosis. Eur Respir J 2017;49.

18. Rybak A, Levy C, Angoulvant F, et al. Association of nonpharmaceutical interventions during the COVID-19 pandemic with invasive pneumococcal disease, pneumococcal carriage, and respiratory viral infections among children in France. JAMA Netw Open 2022;5:e2218959.

19. Danino D, Ben-Shimol S, van der Beek BA, et al. Decline in Pneumococcal Disease in Young Children During the Coronavirus Disease 2019 (COVID-19) Pandemic in Israel Associated With Suppression of Seasonal Respiratory Viruses, Despite Persistent Pneumococcal Carriage: A Prospective Cohort Study. Clin Infect Dis 2022;75:e1154–64.

20. Wildenbeest JG, Zuurbier RP, Korsten K, et al. Respiratory Syncytial Virus Consortium in Europe (RESCEU) birth cohort study: defining the burden of infant respiratory syncytial virus disease in Europe. J Infect Dis 2020;222:S606–S12.

21. Jefferies K, Drysdale SB, Robinson H, et al. Presumed risk factors and biomarkers for severe respiratory syncytial virus disease and related sequelae: protocol for an observational multicenter, case-control study from the respiratory syncytial virus consortium in Europe (RESCEU). J Infect Dis 2020;222:S658–S65.

22. Justicia-Grande AJ, Pardo-Seco J, Cebey-Lopez M, et al. Development and validation of a new clinical scale for infants with acute respiratory infection: the ReSVinet scale. PLoS One 2016;11:e0157665.

23. Lin GL, Golubchik T, Drysdale S, et al. Simultaneous viral whole-genome sequencing and differential expression profiling in respiratory syncytial virus infection of infants. J Infect Dis 2020;222:S666–S71.

24. Lin GL, Drysdale SB, Snape MD, et al. Distinct patterns of within-host virus populations between two subgroups of human respiratory syncytial virus. Nat Commun 2021;12:5125.

25. McGinley JP, Lin GL, Öner D, et al. Clinical and viral factors associated with disease severity and subsequent wheezing in infants with respiratory syncytial virus infection. J Infect Dis 2022;226:S45–54.

26. Bonsall D, Golubchik T, de Cesare M, et al. A comprehensive genomics solution for HIV surveillance and clinical monitoring in low income settings. J Clin Microbiol 2020;58:e00382–20.

27. Bonsall D, Ansari MA, Ip C, et al. ve-SEQ: robust, unbiased enrichment for streamlined detection and whole-genome sequencing of HCV and other highly diverse pathogens. F1000Res 2015;4:1062.

28. Goh C, Golubchik T, Ansari MA, et al. Targeted metagenomic sequencing enhances the identification of pathogens associated with acute infection. bioRxiv 2019:716902.

29. Korsten K, Adriaenssens N, Coenen S, et al. Burden of respiratory syncytial virus infection in community-dwelling older adults in Europe (RESCEU): an international prospective cohort study. Eur Respir J 2021;57:2002688.

30. Wymant C, Blanquart F, Golubchik T, et al. Easy and accurate reconstruction of whole HIV genomes from short-read sequence data with shiver. Virus Evol 2018;4:vey007.

31. Bolger AM, Lohse M, Usadel B. Trimmomatic: a flexible trimmer for Illumina sequence data. Bioinformatics 2014;30:2114–20.

32. Hunt M, Gall A, Ong SH, et al. IVA: accurate de novo assembly of RNA virus genomes. Bioinformatics 2015;31:2374–6.

33. Nurk S, Meleshko D, Korobeynikov A, Pevzner PA. metaSPAdes: a new versatile metagenomic assembler. Genome Res 2017;27:824–34.

34. Langmead B, Salzberg SL. Fast gapped-read alignment with Bowtie 2. Nat Methods 2012;9:357–9.

35. Katoh K, Standley DM. MAFFT multiple sequence alignment software version 7: improvements in performance and usability. Mol Biol Evol 2013;30:772–80.

36. Gotoh O. Optimal alignment between groups of sequences and its application to multiple sequence alignment. Comput Appl Biosci 1993;9:361–70.

37. Stamatakis A. RAxML version 8: a tool for phylogenetic analysis and post-analysis of large phylogenies. Bioinformatics 2014;30:1312–3.

38. Yu G, Smith DK, Zhu H, Guan Y, Lam TTY. ggtree: an R package for visualization and annotation of phylogenetic trees with their covariates and other associated data. Methods Ecol Evol 2017;8:28–36.

39. Lythgoe KA, Hall M, Ferretti L, et al. SARS-CoV-2 within-host diversity and transmission. Science 2021;372:eabg0821.

40. Kroneman A, Vennema H, Deforche K, et al. An automated genotyping tool for enteroviruses and noroviruses. J Clin Virol 2011;51:121–5.

41. Jolley KA, Bliss CM, Bennett JS, et al. Ribosomal multilocus sequence typing: universal characterization of bacteria from domain to strain. Microbiology (Reading) 2012;158:1005–15.

42. Jolley KA, Bray JE, Maiden MCJ. Open-access bacterial population genomics: BIGSdb software, the PubMLST.org website and their applications. Wellcome Open Res 2018;3:124.

43. Jolley KA, Bray JE, Maiden MCJ. A RESTful application programming interface for the PubMLST molecular typing and genome databases. Database (Oxford) 2017;2017:bax060.

44. Wood DE, Lu J, Langmead B. Improved metagenomic analysis with Kraken 2. Genome Biol 2019;20:257.

45. Ondov BD, Bergman NH, Phillippy AM. Interactive metagenomic visualization in a Web browser. BMC Bioinformatics 2011;12:385.

46. Fritz SA, Purvis A. Selectivity in mammalian extinction risk and threat types: a new measure of phylogenetic signal strength in binary traits. Conserv Biol 2010;24:1042–51.

47. Cohen J. Statistical power analysis for the behavioral sciences. 2nd ed: Hillsdale, N.J.; Hove: Erlbaum Associates; 1988.

48. R Core Team. R: a language and environment for statistical computing. Vienna, Austria: R Foundation for Statistical Computing; 2018.

49. Aswad A, Aimola G, Wight D, et al. Evolutionary history of endogenous human herpesvirus 6 reflects human migration out of Africa. Mol Biol Evol 2021;38:96–107.

50. Jain S, Williams DJ, Arnold SR, et al. Community-acquired pneumonia requiring hospitalization among U.S. children. N Engl J Med 2015;372:835–45.

51. Mação P, Dias A, Azevedo L, Jorge A, Rodrigues C. Acute bronchiolitis: a prospective study. Acta Med Port 2011;24 Suppl 2:407–12.

52. Franz A, Adams O, Willems R, et al. Correlation of viral load of respiratory pathogens and co-infections with disease severity in children hospitalized for lower respiratory tract infection. J Clin Virol 2010;48:239–45.

53. Renois F, Talmud D, Huguenin A, et al. Rapid detection of respiratory tract viral infections and coinfections in patients with influenza-like illnesses by use of reverse transcription-PCR DNA microarray systems. J Clin Microbiol 2010;48:3836–42.

54. Pigny F, Wagner N, Rohr M, et al. Viral co-infections among SARS-CoV-2-infected children and infected adult household contacts. Eur J Pediatr 2021;180:1991–5.

55. Jain S, Self WH, Wunderink RG, et al. Community-acquired pneumonia requiring hospitalization among U.S. adults. N Engl J Med 2015;373:415–27.

56. Achten NB, Wu P, Bont L, et al. Interference between respiratory syncytial virus and human rhinovirus infection in infancy. J Infect Dis 2017;215:1102–6.

57. Karppinen S, Toivonen L, Schuez-Havupalo L, Waris M, Peltola V. Interference between respiratory syncytial virus and rhinovirus in respiratory tract infections in children. Clin Microbiol Infect 2016;22:208.e1-.e6.

58. de Steenhuijsen Piters WA, Heinonen S, Hasrat R, et al. Nasopharyngeal microbiota, host transcriptome, and disease severity in children with respiratory syncytial virus infection. Am J Respir Crit Care Med 2016;194:1104–15.

59. Man WH, van Houten MA, Mérelle ME, et al. Bacterial and viral respiratory tract microbiota and host characteristics in children with lower respiratory tract infections: a matched case-control study. Lancet Respir Med 2019;7:417–26.

60. de Koff EM, Man WH, van Houten MA, et al. Microbial and clinical factors are related to recurrence of symptoms after childhood lower respiratory tract infection. ERJ Open Res 2021;7:00939–2020.

61. Sleeman KL, Daniels L, Gardiner M, et al. Acquisition of Streptococcus pneumoniae and nonspecific morbidity in infants and their families: a cohort study. Pediatr Infect Dis J 2005;24:121–7.

62. Ederveen THA, Ferwerda G, Ahout IM, et al. Haemophilus is overrepresented in the nasopharynx of infants hospitalized with RSV infection and associated with increased viral load and enhanced mucosal CXCL8 responses. Microbiome 2018;6:10.

63. Gulraiz F, Bellinghausen C, Bruggeman CA, Stassen FR. Haemophilus influenzae increases the susceptibility and inflammatory response of airway epithelial cells to viral infections. FASEB J 2015;29:849–58.

64. Coultas JA, Smyth R, Openshaw PJ. Respiratory syncytial virus (RSV): a scourge from infancy to old age. Thorax 2019;74:986–93.

