## Supplemental tables and figures for "Association between disease severity and co-detection of respiratory pathogens in infants with RSV infection"

Table S1. Target bacterial and viral species.

| **Bacteria** | | |
|---|---|---|
| *Acinetobacter baumannii* | *Escherichia coli* | *Neisseria meningitidis* |
| *Acinetobacter calcoaceticus* | *Haemophilus influenzae* | *Pseudomonas aeruginosa* |
| *Bartonella henselae* | *Haemophilus parainfluenzae* | *Serratia marcescens* |
| *Bordetella pertussis* | *Klebsiella pneumoniae* | *Staphylococcus aureus* |
| *Borrelia burgdorferi* | *Klebsiella oxytoca* | *Stenotrophomonas maltophilia* |
| *Brucella* spp. | *Legionella pneumophila* | *Streptococcus agalactiae* |
| *Chlamydia psittaci* | *Leptospira* spp. | *Streptococcus pneumoniae* |
| *Chlamydophila pneumoniae* | *Listeria monocytogenes* | *Streptococcus pyogenes* |
| *Coxiella burnetiid* | *Moraxella catarrhalis* | *Treponema pallidum* |
| *Enterobacter aerogenes* | *Mycobacterium tuberculosis* | |
| *Enterobacter cloacae* | *Mycoplasma pneumoniae* | |

| **Viruses** | |
|---|---|
| BK polyomavirus | Lassa mammarenavirus |
| California encephalitis virus | Lymphocytic chriomeningitis mammarenavirus |
| Cardiovirus A & B | Mastadenovirus A–G |
| Coronavirus HCoV-229E | Measles virus A, B1–B3, C1, C2, D1–D11, E, F, G1–G3, H1, H2 |
| Coronavirus HCoV-HKU1 genotypes A, B, C | Mumps virus A–D, F–L, N |
| Coronavirus HCoV-NL63 | Murray Valley encephalitis virus (all genotypes) |
| Coronavirus HCoV-OC43 genotypes A-E | Primate erythroparvovirus 1 |
| Coronavirus MERS | Primate tetraparvovirus 1 |
| Coronavirus SARS | Respiratory syncytial virus A & B |
| Dengue virus 1–4 | Rhabdovirus 1—Rabies virus |
| Eastern equine encephalitis virus | Rhabdovirus 2—Lagos bat virus |

| | |
|---|---|
| Enterovirus A & B | Rhabdovirus 3—Mokola virus |
| Hendra virus | Rhabdovirus 4—Duvenhage virus |
| Henipavirus B & M | Rhabdovirus 5—European bat lyssavirus 1 |
| Hepatitis A virus | Rhabdovirus 6—European bat lyssavirus 2 |
| HHV1—Herpes simplex virus type 1[a] | Rhabdovirus 7—Australian bat lyssaviruses |
| HHV2—Herpes simplex virus type 2[a] | Rhinovirus A–C |
| HHV3—Varicella-zoster virus[a] | Rift Valley fever virus |
| HHV4—Epstein-Barr virus[a] | Rosavirus A |
| HHV5—Human cytomegalovirus[a] | Rotavirus A–C |
| HHV6A—Human herpesvirus 6A[a] | Rubella virus |
| HHV6B—Human herpesvirus 6B[a] | Salivirus A & FHB |
| HHV7—Human herpesvirus 7[a] | Sandfly fever Naples virus |
| HHV8—Kaposi's sarcoma herpesvirus[a] | Sandfly fever Sicilian virus |
| Human bocavirus 1 | Sosuga virus |
| Human metapneumovirus | St. Louis encephalitis virus (all genotypes) |
| Human parainfluenza virus 1, 2, 3, 4a, 4b, 5 | Tick-borne encephalitis virus (all genotypes) |
| Human parechovirus A and B | Venezuelan equine encephalitis virus |
| Influenza A virus H1N1, H1N2, H2N2, H3N2, H5N1, H7N3, H7N7, H7N9, H9N2 | West Nile virus (all genotypes) |
| Influenza B virus | Western equine encephalitis virus |
| Influenza C virus | Yellow fever virus (all genotypes) |
| Japanese encephalitis virus (all genotypes) | Zika virus |
| JC polyomavirus | |

[a] HHV denotes human herpesvirus; the probe set only targets partial genomes of the HHVs.

Table S2. Criteria for the presence and absence of each target virus.

| Virus | Criteria[a] |
|---|---|
| Human adenovirus | Presence: >50 unique reads & >30% coverage<br>Absence: <50 unique reads & <10% coverage |
| Human coronavirus | Presence: >50 unique reads<br>Absence: <50 unique reads & <10% coverage |
| Human cytomegalovirus | Presence: >30 unique reads or >30% coverage<br>Absence: <10 unique reads |
| Enterovirus | Presence: >20 unique reads or >40% coverage<br>Absence: <10 unique reads & ≤40% coverage |
| Human herpesvirus 6 | Presence: >50 unique reads<br>Absence: 0 read |
| Influenza C virus | Presence: >50 unique reads<br>Absence: 0 read |
| Human parainfluenza virus | Presence: >20 unique reads<br>Absence: <5 unique reads |
| Human parechovirus A | Presence: >25 unique reads<br>Absence: ≤10 pre-deduplicated reads |
| Human bocavirus | Presence: >50 unique reads<br>Absence: <10 unique reads |

[a] Coverage represents the percentage of genomic regions covered by a minimum of 2 reads (without deduplication).

Table S3. Number of RSV-infected infants from each country and each season (N = 440).

| | Longitudinal birth cohort study (N = 146) | Infant cross-sectional study (N = 294) |
|---|---|---|
| 2017–18 | 13 | 57 |
| Spain | 10 | 0 |
| United Kingdom | 0 | 6 |
| Netherlands | 3 | 51 |
| 2018–19 | 41 | 122 |
| Spain | 10 | 10 |
| United Kingdom | 12 | 75 |
| Netherlands | 19 | 37 |
| 2019–20 | 92 | 115 |
| Spain | 32 | 15 |
| United Kingdom | 18 | 58 |
| Netherlands | 42 | 42 |

Table S4. Demographic and clinical characteristics of RSV-infected infants, stratified by age (N = 431).[a]

| | <3 mo (N = 169) | 3 to <6 mo (N = 117) | 6 to <12 mo (N = 145) | P value |
|---|---|---|---|---|
| **Demographic features** | | | | |
| Gestational age | | | | |
| Median (IQR) — wk | 39.3 (38.0–40.1) | 40.0 (39.0–40.7) | 39.7 (38.7–40.7) | $7.3\times10^{-4}$ |
| Distribution | | | | 0.399 |
| <32 wk | 4/168 (2) | 1/115 (1) | 3/144 (2) | |
| 32 to <37 wk | 10/168 (6) | 2/115 (2) | 5/144 (3) | |
| ≥37 wk | 154/168 (92) | 112/115 (97) | 136/144 (94) | |
| Female sex | 66 (39) | 51 (44) | 75 (52) | 0.077 |
| Comorbidity | 18 (11) | 5 (4) | 14 (10) | 0.142 |
| **Virological features** | | | | |
| RSV-A[b] | 79/165 (48) | 66/114 (58) | 74/143 (52) | 0.442 |
| Peak RSV read count — total no. | 166 | 114 | 141 | |
| Mean ± SD — $\log_{10}$ | 3.8 ± 1.1 | 4.2 ± 1.0 | 4.1 ± 1.0 | 0.081[c] |
| **Clinical features**[d] | | | | |
| ReSVinet score | | | | |
| Mean ± SD | 9.6 ± 4.9 | 6.7 ± 4.5 | 5.9 ± 3.3 | $2.4\times10^{-6}$ |
| Distribution | | | | $5.7\times10^{-5}$ |
| 0–7 | 63/166 (38) | 74/113 (65) | 102/140 (73) | |
| 8–13 | 60/166 (36) | 28/113 (25) | 34/140 (24) | |
| 14–20 | 43/166 (26) | 11/113 (10) | 4/140 (3) | |
| Fever | 33/166 (20) | 39/113 (35) | 70/140 (50) | $3.5\times10^{-8}$ |
| Hospitalisation | 130/167 (78) | 47/104 (45) | 42/132 (32) | $9.6\times10^{-11}$ |
| PICU admission | 57/167 (34) | 15/104 (14) | 5/132 (4) | $3.7\times10^{-6}$ |
| Respiratory support | 116/157 (74) | 36/92 (39) | 27/122 (22) | $6.4\times10^{-13}$ |
| Mechanical ventilation | 53/157 (34) | 10/92 (11) | 3/122 (2) | $8.7\times10^{-7}$ |

[a] Two participants without available age information were excluded from this table. IQR denotes interquartile range; SD, standard deviation; PICU, paediatric intensive care unit. Unless otherwise specified, data are shown as number/total number (%) or number (%) if there is no missing value. Percentages may not total 100 due to rounding. For demographic features, Kruskal–Wallis tests were used to compare continuous variables between the groups; chi-square tests with Yates' correction or Fisher's exact tests were used to compare categorical variables between the groups, whichever is appropriate.

[b] Nine participants with both RSV subgroups A and B identified were excluded from this comparison. Multivariable logistic regression was used to adjust for the sampling season.

[c] Multiple linear regression was used to adjust for the duration between symptom onset and sampling.

[d] Multiple linear regression, ordered logistic regression, or multivariable logistic regression was used to adjust for covariates, depending on the type of the response (dependent) variable. Covariates included gestational age, sex, comorbidity, sampling season and country, study, RSV subgroup, peak RSV read count, and the duration between symptom onset and sampling. Models with different combinations of the covariates were tested, and the model with the lowest Akaike information criterion (AIC) was selected. Likelihood-ratio tests were used to assess the effect of age on the goodness of fit of these models.

Table S5. Demographic and clinical characteristics of RSV-infected infants, stratified by study (N = 433).[a]

| | Longitudinal birth cohort study (N = 143) | Infant cross-sectional study (N = 290) | P value |
|---|---|---|---|
| **Demographic features** | | | |
| Age | | | |
| Median (IQR) — mo | 5.7 (3.6–8.9) | 3.0 (1.5–6.4) | $2.5\times10^{-10}$ |
| Distribution — no./total no. (%) | | | $8.3\times10^{-10}$ |
| <3 mo | 25/142 (18) | 144/289 (50) | |
| 3 to <6 mo | 50/142 (35) | 67/289 (23) | |
| 6 to <12 mo | 67/142 (47) | 78/289 (27) | |
| Gestational age | | | |
| Median (IQR) — wk | 39.9 (39.0–40.9) | 39.4 (38.0–40.3) | $9.8\times10^{-5}$ |
| Distribution — no./total no. (%) | | | $3.3\times10^{-4}$ |
| <32 wk | 0/139 (0) | 8/288 (3) | |
| ≥32 to <37 wk | 0/139 (0) | 17/288 (6) | |
| ≥37 wk | 139/139 (100) | 263/288 (91) | |
| Female sex — no./total no. (%) | 67/142 (47) | 125/289 (43) | 0.504 |
| Comorbidity — no./total no. (%) | 0/143 (0) | 37/289 (13) | $1.9\times10^{-7}$ |
| **Virological features** | | | |
| RSV-A — no./total no. (%)[b] | 75/142 (53) | 145/282 (51) | 0.207 |
| Peak RSV read count — total no. | 136 | 286 | |
| Mean ± SD — $\log_{10}$ | 4.2 ± 1.0 | 3.9 ± 1.0 | 0.079[c] |
| **Clinical features**[d] | | | |
| ReSVinet score | | | |
| Mean ± SD | 4.7 ± 2.6 | 8.9 ± 4.7 | $2.6\times10^{-9}$ |
| Distribution — no./total no. (%) | | | $7.5\times10^{-9}$ |
| 0–7 | 120/136 (88) | 120/284 (42) | |
| 8–13 | 15/136 (11) | 107/284 (38) | |
| 14–20 | 1/136 (1) | 57/284 (20) | |

| | | | |
|---|---|---|---|
| Fever — no./total no. (%) | 53/136 (39) | 89/284 (31) | 0.334 |
| Hospitalisation — no./total no. (%) | 7/115 (6) | 212/289 (73) | $3.9 \times 10^{-12}$ |
| PICU admission — no./total no. (%) | 1/115 (1) | 76/289 (26) | 0.002 |
| Respiratory support — no./total no. (%) | 3/112 (3) | 176/260 (68) | $8.5 \times 10^{-11}$ |
| Mechanical ventilation — no./total no. (%) | 0/112 (0) | 66/260 (25) | 0.985 |

[a] IQR denotes interquartile range; SD, standard deviation; PICU, paediatric intensive care unit. For demographic features, Mann–Whitney U tests were used to compare continuous variables between the two groups; chi-square tests with Yates' correction or Fisher's exact tests were used to compare categorical variables between the two groups, whichever is appropriate.

[b] Nine participants with both RSV subgroups A and B identified were excluded from this comparison. Multivariable logistic regression was used to adjust for the sampling season.

[c] Multiple linear regression was used to adjust for the duration between symptom onset and sampling.

[d] Multiple linear regression, ordered logistic regression, or multivariable logistic regression was used to adjust for covariates, depending on the type of the response (dependent) variable. Covariates included age, gestational age, sex, comorbidity, RSV subgroup, sampling season and country, peak RSV read count, and the duration between symptom onset and sampling. Models with different combinations of the covariates were tested, and the model with the lowest Akaike information criterion (AIC) was selected.

Table S6. Incidences of the two RSV subgroups in each country and season (N = 433).

| | RSV-A (N = 220) | RSV-B (N = 204) | Mixed (N = 9) |
|---|---|---|---|
| 2017–18 | 19 | 50 | 1 |
| Spain | 0 | 10 | 0 |
| United Kingdom | 4 | 2 | 0 |
| Netherlands | 15 | 38 | 1 |
| 2018–19 | 68 | 89 | 4 |
| Spain | 13 | 7 | 0 |
| United Kingdom | 40 | 45 | 2 |
| Netherlands | 15 | 37 | 2 |
| 2019–20 | 133 | 65 | 4 |
| Spain | 29 | 18 | 0 |
| United Kingdom | 45 | 29 | 0 |
| Netherlands | 59 | 18 | 4 |

Table S7. Characteristics of the RSV-infected infants by RSV subgroup (N = 424).[a]

| | RSV-A (N = 220) | RSV-B (N = 204) | P value | Q value |
|---|---|---|---|---|
| **Demographic features** | | | | |
| Age | | | | |
| Median (IQR) — mo | 4.3 (1.9–7.3) | 3.8 (1.7–7.5) | 0.446 | |
| Distribution — no./total no. (%) | | | 0.258 | |
| <3 mo | 79/219 (36) | 86/203 (42) | | |
| 3 to <6 mo | 66/219 (30) | 48/203 (24) | | |
| 6 to <12 mo | 74/219 (34) | 69/203 (34) | | |
| Gestational age | | | | |
| Median (interquartile range) — wk | 39.9 (38.5–40.6) | 39.6 (38.6–40.3) | 0.501 | |
| Distribution — no./total no. (%) | | | 0.508 | |
| <32 wk | 3/215 (1) | 5/203 (2) | | |
| 32 to <37 wk | 10/215 (5) | 6/203 (3) | | |
| ≥37 wk | 202/215 (94) | 192/203 (95) | | |
| Female sex — no./total no. (%) | 95/219 (43) | 93/203 (46) | 0.686 | |
| Comorbidity — no./total no. (%) | 16/220 (7) | 20/203 (10) | 0.438 | |
| **Virological features** | | | | |
| Peak RSV read count — total no. | 214 | 199 | | |
| Mean ± SD — $\log_{10}$ | 4.1 ± 0.9 | 3.9 ± 1.1 | 0.067[b] | |
| **Clinical features**[c] | | | | |
| ReSVinet score | | | | |
| Mean ± SD | 7.4 ± 4.2 | 7.6 ± 5.0 | 0.457 | 0.639 |
| Distribution — no./total no. (%) | | | 0.662 | 0.761 |
| 0–7 | 127/216 (59) | 110/195 (56) | | |
| 8–13 | 64/216 (30) | 54/195 (28) | | |
| 14–20 | 25/216 (12) | 31/195 (16) | | |
| Fever — no./total no. (%) | 80/216 (37) | 58/195 (30) | 0.040 | 0.140 |
| Hospitalisation — no./total no. (%) | 115/205 (56) | 97/190 (51) | 0.034 | 0.140 |

| | | | | |
|---|---|---|---|---|
| PICU admission — no./total no. (%) | 31/205 (15) | 42/190 (22) | 0.346 | 0.605 |
| Any respiratory support — no./total no. (%) | 89/186 (48) | 84/178 (47) | 0.330 | 0.605 |
| Mechanical ventilation — no./total no. (%) | 27/186 (15) | 36/178 (20) | 0.761 | 0.761 |

[a] Nine participants with both RSV subgroups A and B identified were excluded from this table. IQR denotes interquartile range; SD, standard deviation; PICU, paediatric intensive care unit. Percentages may not total 100 due to rounding. For demographic features, Mann–Whitney U tests were used to compare continuous variables between the two groups; chi-square tests with Yates' correction or Fisher's exact tests were used to compare categorical variables between the two groups, whichever is appropriate.

[b] Multiple linear regression was used to adjust for the duration between symptom onset and sampling.

[c] Multiple linear regression, ordered logistic regression, or multivariable logistic regression was used to adjust for covariates, depending on the type of the response (dependent) variable. Covariates included age, gestational age, sex, comorbidity, sampling season and country, study, peak RSV read count, and the duration between symptom onset and sampling. Models with different combinations of the covariates were tested, and the model with the lowest Akaike information criterion (AIC) was selected.

Table S8. Demographic and clinical features of the RSV-infected infants with and without co-detection of any other virus (N = 405).[a]

| | Presence of any other virus (N = 106) | Absence of any other virus (N = 299) | P value | Q value |
|---|---|---|---|---|
| **Demographic features** | | | | |
| Age | | | | |
| Median (IQR) — mo | 4.3 (2.3–8.2) | 3.7 (1.7–6.9) | 0.031 | |
| Distribution — no./total no. (%) | | | 0.203 | |
| <3 mo | 35/105 (33) | 129/298 (43) | | |
| 3 to <6 mo | 31/105 (30) | 75/298 (25) | | |
| 6 to <12 mo | 39/105 (37) | 94/298 (32) | | |
| Female sex — no./total no. (%) | 43/105 (41) | 132/298 (44) | 0.631 | |
| Comorbidity — no./total no. (%) | 15/105 (14) | 21/299 (7) | 0.041 | |
| **Virological features** | | | | |
| RSV-A — no./total no. (%)[b] | 55/104 (53) | 149/292 (51) | 0.680 | |
| Peak RSV read count — total no. | 103 | 291 | | |
| Mean ± SD — $\log_{10}$ | 3.8 ± 1.1 | 4.1 ± 1.0 | 0.107[c] | |
| **Clinical features**[d] | | | | |
| ReSViNET score | | | | |
| Mean ± SD | 7.9 ± 5.1 | 7.7 ± 4.5 | 0.203 | 0.284 |
| Distribution — no./total no. (%) | | | 0.066 | 0.115 |
| 0–7 | 54/104 (52) | 164/289 (57) | | |
| 8–13 | 33/104 (32) | 85/289 (29) | | |
| 14–20 | 17/104 (16) | 40/289 (14) | | |
| Fever — no./total no. (%) | 34/104 (33) | 97/289 (34) | 0.276 | 0.322 |
| Hospitalisation — no./total no. (%) | 52/96 (54) | 157/285 (55) | 0.541 | 0.541 |
| PICU admission — no./total no. (%) | 25/96 (26) | 51/285 (18) | 0.005 | 0.025 |
| Any respiratory support — no./total no. (%) | 46/87 (53) | 127/264 (48) | 0.027 | 0.063 |
| Mechanical ventilation — no./total no. (%) | 21/87 (24) | 44/264 (17) | 0.007 | 0.025 |

[a] Twenty-eight patients with an equivocal presence of any other virus were removed from this table. IQR denotes interquartile range; SD, standard deviation; PICU, paediatric intensive care unit. For demographic features, Mann–Whitney U tests were used to compare continuous variables between the two groups, and chi-square tests with Yates' correction were used to compare categorical variables between the two groups.

[b] Nine participants with both RSV subgroups A and B identified were excluded from this comparison. Multivariable logistic regression was used to adjust for the sampling season.

[c] Multiple linear regression was used to adjust for the duration between symptom onset and sampling.

[d] Multiple linear regression, ordered logistic regression, or multivariable logistic regression was used to adjust for covariates, depending on the type of the response (dependent) variable. Covariates included age, gestational age, sex, comorbidity, sampling season and country, study, RSV subgroup, peak RSV read count, and the duration between symptom onset and sampling. Models with different combinations of the covariates were tested, and the model with the lowest Akaike information criterion (AIC) was selected.

Table S9. The presence and absence of viruses additional to RSV in samples from RSV-infected infants (N = 433).[a]

| | Presence | Absence | Equivocal |
|---|---|---|---|
| Any non-RSV virus | 106 (24) | 299 (69) | 28 (6) |
| Enterovirus (any) | 69 (16) | 355 (82) | 9 (2) |
| – Rhinovirus | 63 (15) | 362 (84) | 8 (2) |
| – Enterovirus D | 3 (1) | 429 (99) | 1 (<1) |
| – Enterovirus A | 2 (<1) | 431 (100) | 0 |
| – Enterovirus B | 1 (<1) | 431 (100) | 1 (<1) |
| Human coronavirus (any)[b] | 16 (4) | 415 (96) | 2 (<1) |
| – OC43 | 7 (2) | 425 (98) | 1 (<1) |
| – HKU1 | 6 (1) | 427 (99) | 0 |
| – NL63 | 2 (<1) | 430 (99) | 1 (<1) |
| – 229E | 1 (<1) | 432 (100) | 0 |
| Human adenovirus | 10 (2) | 419 (97) | 4 (1) |
| Human herpesvirus 6 | 7 (2) | 414 (96) | 12 (3) |
| Human bocavirus | 4 (1) | 428 (99) | 1 (<1) |
| Human parainfluenza virus | 3 (1) | 429 (99) | 1 (<1) |
| Human cytomegalovirus | 3 (1) | 426 (98) | 4 (1) |
| Human parechovirus A | 3 (1) | 425 (98) | 5 (1) |
| Influenza C virus | 1 (<1) | 432 (100) | 0 |

[a] Data are shown as number (percentage). Percentages may not total 100 due to rounding.

[b] Some samples were collected in late 2019 and early 2020 (after the emergence of SARS-CoV-2).

Table S10. Phylogenetic signals between the RSV phylogenies and commonly co-detected pathogens.[a]

A. RSV-A

| | HRV | HCoV | AdV | HHV-6 |
|---|---|---|---|---|
| *D* value | 0.934 | 1.054 | 1.328 | — |
| Probability of random distribution of the virus | 0.274 | 0.581 | 0.783 | — |
| Probability of distribution of the virus under Brownian motion | 0 | 0.003 | 0 | — |
| | *M. catarrhalis* | *S. pneumoniae* | *H. influenzae* | *S. aureus* |
| *D* value | 0.994 | 0.989 | 0.990 | 0.938 |
| Probability of random distribution of the bacterium | 0.459 | 0.434 | 0.439 | 0.350 |
| Probability of distribution of the bacterium under Brownian motion | 0 | 0 | 0 | 0 |

B. RSV-B

| | HRV | HCoV | AdV | HHV-6 |
|---|---|---|---|---|
| *D* value | 1.171 | 0.802 | — | 0.975 |
| Probability of random distribution of the virus | 0.813 | 0.255 | — | 0.531 |
| Probability of distribution of the virus under Brownian motion | 0 | 0.012 | — | 0.047 |
| | *M. catarrhalis* | *S. pneumoniae* | *H. influenzae* | *S. aureus* |
| *D* value | 1.009 | 0.772 | 0.917 | 0.759 |
| Probability of random distribution of the bacterium | 0.508 | 0.026 | 0.240 | 0.146 |
| Probability of distribution of the bacterium under Brownian motion | 0 | 0 | 0 | 0 |

[a] Viruses that were co-detected with less than three RSV strains on the phylogeny were not tested for phylogenetic signal (see Fig. S6). HRV denotes human rhinovirus; HCoV, human coronavirus; AdV, adenovirus; HHV-6, human herpesvirus 6; *M. catarrhalis*, *Moraxella catarrhalis*; *S. pneumoniae*, *Streptococcus pneumoniae*; *H. influenzae*, *Haemophilus influenzae*; *S. aureus*, *Staphylococcus aureus*.

Table S11. The presence and absence of each identified bacterial genus in the RSV-infected infants (N = 433).[a]

| | Presence | Absence | Equivocal |
|---|---|---|---|
| *Moraxella* | 327 (76) | 99 (23) | 7 (2) |
| – *M. catarrhalis* | 286 (66) | 139 (32) | 8 (2) |
| *Streptococcus* | 296 (68) | 76 (18) | 61 (14) |
| – *S. pneumoniae* | 222 (51) | 187 (43) | 24 (6) |
| *Haemophilus* | 194 (45) | 225 (52) | 14 (3) |
| – *H. influenzae* | 164 (38) | 262 (61) | 7 (2) |
| *Neisseria* | 50 (12) | 362 (84) | 21 (5) |
| – *N. meningitidis* | 2 (<1) | 431 (>99) | 0 |
| *Staphylococcus* | 49 (11) | 372 (86) | 12 (3) |
| – *S. aureus* | 38 (9) | 388 (90) | 7 (2) |
| *Burkholderia*[b] | 37 (9) | 362 (84) | 34 (8) |
| *Dolosigranulum*[b] | 34 (8) | 382 (88) | 17 (4) |
| *Gemella*[b] | 19 (4) | 400 (92) | 14 (3) |
| *Enterococcus*[b] | 16 (4) | 406 (94) | 11 (3) |
| *Granulicatella*[b] | 8 (2) | 417 (96) | 8 (2) |
| *Rothia*[b] | 7 (2) | 422 (97) | 4 (1) |
| *Enterobacter* | 5 (1) | 427 (99) | 1 (<1) |
| *Escherichia* | 5 (1) | 427 (99) | 1 (<1) |
| *Corynebacterium*[b] | 5 (1) | 419 (97) | 9 (2) |
| *Pasteurella*[b] | 3 (1) | 425 (98) | 5 (1) |
| *Veillonella*[b] | 3 (1) | 423 (98) | 7 (2) |
| *Klebsiella* | 2 (<1) | 430 (99) | 1 (<1) |
| *Aggregatibacter*[b] | 2 (<1) | 429 (99) | 2 (<1) |
| *Citrobacter*[b] | 1 (<1) | 432 (>99) | 0 |
| *Xanthomonas*[b] | 1 (<1) | 432 (>99) | 0 |
| *Lactococcus*[b] | 1 (<1) | 431 (>99) | 1 (<1) |
| *Acinetobacter* | 1 (<1) | 430 (99) | 2 (<1) |
| *Actinomyces*[b] | 1 (<1) | 429 (99) | 3 (1) |
| *Stenotrophomonas* | 1 (<1) | 429 (99) | 3 (1) |
| *Pseudomonas* | 1 (<1) | 422 (97) | 10 (2) |

[a] Data are shown as number (percentage). Percentages may not total 100 due to rounding.

[b] These genera were not included in the *Castanet* enrichment panel, so their read numbers were expected to be lower as these were detected by unenriched metagenomics only.

Table S12. Frequency of known ribosomal sequence types of the bacteria recovered from this study and the most likely corresponding sequence type and serotype combination.

| Frequency | Ribosomal sequence type | Sequence type[a] | Clonal complex[a] | Serotype[b] | %[c] | Total # of isolates in PubMLST[d] |
|---|---|---|---|---|---|---|
| *Streptococcus pneumoniae* | | | | | | |
| 1 | 614 | 138 | - | 6B | 75 | 73 |
| 2 | 644 | 62 | - | 11A | 59 | 145 |
| 2 | 12187 | 2062 | - | 19A | 84 | 90 |
| 1 | 12227 | 66 | - | 9N | 73 | 60 |
| 1 | 12688 | 30 | - | Inconclusive | 80 | 10 |
| 1 | 13052 | 1925 | - | 19A | 100 | 4 |
| 1 | 13435 | 439 | - | 23B | 56 | 155 |
| 2 | 13906 | 1262 | - | 15BC | 62 | 166 |
| 1 | 21764 | 392 | - | 17F | 56 | 9 |
| 1 | 22747 | 1349 | - | Genetic variant | 42 | 12 |
| 1 | 22892 | 4149 | - | Inconclusive | 81 | 16 |
| 1 | 102491 | 2105 | - | 15A | 38 | 8 |
| 2 | 104663 | 3811 | - | 15A | 100 | 12 |
| 1 | 151444 | 452 | - | Inconclusive | 100 | 8 |
| 1 | 151477 | 2669 | - | Inconclusive | 88 | 8 |
| 1 | 151481 | 179 | - | Inconclusive | 100 | 3 |
| *Haemophilus influenzae* | | | | | | |
| 1 | 24137 | 1877 | ST-584 | NT | 31 | 16 |
| 2 | 24162 | 12 | ST-12 | NT | 66 | 44 |
| 1 | 24179 | 422 | ST-422 | NT | 58 | 31 |
| 1 | 24185 | 165 | ST-165 | NT | 76 | 25 |
| 1 | 24208 | 57 | ST-57 | NT | 78 | 18 |
| 2 | 49634 | 105 | ST-105 | NT | 100 | 1 |
| 1 | 66853 | 368 | ST-1836 | NT | 88 | 8 |
| 1 | 66856 | 159 | ST-107 | NT | 89 | 18 |
| 1 | 66889 | 145 | ST-11 | NT | 88 | 16 |
| 1 | 72352 | 266 | ST-266 | NT | 70 | 10 |
| 2 | 89110 | 160 | ST-487 | NT | 84 | 19 |
| 1 | 89135 | 567 | ST-746 | NT | 75 | 4 |
| 1 | 89141 | 1218 | ST-107 | NT | 93 | 14 |

| Frequency | Ribosomal sequence type | Sequence type[a] | Clonal complex[a] | Serotype[b] | %[c] | Total # of isolates in PubMLST[d] |
|---|---|---|---|---|---|---|
| 1 | 91528 | 932 | - | NT | 100 | 1 |
| 1 | 98191 | 836 | ST-836 | NT | 70 | 10 |
| 1 | 125128 | 2092/2597 | ST-1025 | ND/NT | 50 | 2 |
| 1 | 125166 | 597 | ST-584 | NT | 75 | 4 |
| 1 | 125211 | 3 | ST-3 | NT | 100 | 2 |
| 1 | 126121 | 1238 | ST-931 | NT | 100 | 1 |
| 1 | 126127 | - | - | - | - | - |
| 1 | 132982 | 472 | ST-472 | NT | 60 | 5 |
| 2 | 133371 | - | - | - | - | - |
| *Escherichia coli* | | | | | | |
| 1 | 1674 | 1193/53 | ST-14 | - | 60 | 5 |
| 1 | 2135 | 69/3 | ST-69 | UPEC | 54 | 94 |
| 1 | 93310 | - | - | - | - | - |
| *Neisseria meningitidis* | | | | | | |
| 1 | 2327 | 11 | ST-11 | W | 87 | 2279 |
| *Moraxella catarrhalis* | | | | | | |
| 1 | 49012 | - | - | - | - | - |
| 1 | 89989 | - | - | - | - | - |
| 1 | 92879 | - | - | - | - | - |
| 1 | 92972 | - | - | - | - | - |
| 2 | 105842 | - | - | - | - | - |
| 1 | 131792 | - | - | - | - | - |
| 1 | 131807 | - | - | - | - | - |

ND denotes not determined; NT, nontypeable; UPEC, uropathogenic *E. coli*.

[a] Sequence type and clonal complex are based on the multi-locus sequence typing (MLST) scheme. For *E. coli*, sequence type based on the Achtman MLST scheme is shown first, followed by sequence type based on the Pasteur MLST scheme.

[b] For *E. coli*, phenotype is shown in the serotype column. For *N. meningitidis*, capsule group is shown in the serotype column.

[c] Percentage of this most frequent combination of the ribosomal sequence type, sequence type, clonal complex, and serotype.

[d] Total number of isolates with this specified sequence type found in PubMLST (https://pubmlst.org/organisms) as of 1st December 2022.

Table S13. Demographic and clinical features of the RSV-infected infants with and without co-detection of any *Haemophilus* sp (N = 419).[a]

| | Presence of any *Haemophilus* sp. (N = 194) | Absence of any *Haemophilus* sp. (N = 225) | P value | Q value | Cohen's $f^2$ [b] |
|---|---|---|---|---|---|
| **Demographic features** | | | | | |
| Age | | | | | |
| Median (IQR) — mo | 5.2 (2.0–8.7) | 3.5 (1.7–5.9) | $1.6\times10^{-4}$ | | |
| Distribution | | | $4.2\times10^{-6}$ | | |
| <3 mo | 70/194 (36) | 93/223 (42) | | | |
| 3 to <6 mo | 37/194 (19) | 78/223 (35) | | | |
| 6 to <12 mo | 87/194 (45) | 52/223 (23) | | | |
| Female sex | 84/194 (43) | 101/223 (45) | 0.757 | | |
| Comorbidity | 20/194 (10) | 16/224 (7) | 0.329 | | |
| **Virological features** | | | | | |
| RSV-A[c] | 109/192 (57) | 103/218 (47) | 0.009 | | |
| Peak RSV read count — total no. | 192 | 216 | | | |
| Mean ± SD — $\log_{10}$ | 4.0 ± 0.9 | 4.1 ± 1.1 | 0.745[d] | | |
| **Clinical features**[e] | | | | | |
| ReSVinet score | | | | | |
| Mean ± SD | 8.5 ± 4.5 | 6.8 ± 4.6 | $3.9\times10^{-6}$ | $2.7\times10^{-5}$ | 0.058 |
| Distribution | | | 0.001 | 0.003 | 0.019 |
| 0–7 | 92/191 (48) | 140/215 (65) | | | |
| 8–13 | 68/191 (36) | 49/215 (23) | | | |
| 14–20 | 31/191 (16) | 26/215 (12) | | | |
| Fever | 85/191 (44) | 54/215 (25) | 0.017 | 0.024 | 0.023 |
| Hospitalisation | 112/178 (63) | 99/213 (46) | $4.7\times10^{-5}$ | $1.6\times10^{-4}$ | 0.086 |
| PICU admission | 43/178 (24) | 33/213 (15) | 0.091 | 0.091 | 0.015 |
| Any respiratory support | 90/164 (55) | 83/196 (42) | 0.013 | 0.023 | 0.049 |
| Mechanical ventilation | 40/164 (24) | 25/196 (13) | 0.030 | 0.035 | 0.029 |

[a] Fourteen patients with an equivocal presence of any *Haemophilus* sp. were excluded from the table. *Haemophilus* spp. included *H. influenzae* (80%), *H. haemolyticus* (10%), *H. parainfluenzae* (7%), *H. parahaemolyticus* (1%) and unclassified species (1%). Unless otherwise specified, data are shown as number/total number (%) or number (%) if there is no missing value. Percentages may not total 100 due to rounding. IQR denotes interquartile range; SD, standard deviation; and PICU, paediatric intensive care unit. For demographic features, Mann–Whitney U tests were used to compare continuous variables between the two groups, and chi-square tests with Yates' correction were used to compare categorical variables between the two groups.

[b] Cohen's $f^2$ was used to evaluate the effect size of the presence of any *Haemophilus* sp. on different clinical features. A value of ≥0.02 represents a small effect size; a value of ≥0.15, medium; and a value of ≥0.35, large.

[c] Nine participants with both RSV subgroups A and B identified were excluded from this comparison. Multivariable logistic regression was used to adjust for the sampling season.

[d] Multiple linear regression was used to adjust for the duration between symptom onset and sampling.

[e] Multiple linear regression, ordered logistic regression, or multivariable logistic regression was used to adjust for covariates, depending on the type of the response (dependent) variable. Covariates included age, gestational age, sex, comorbidity, sampling season and country, study, RSV subgroup, peak RSV read count, and the duration between symptom onset and sampling. Models with different combinations of the covariates were tested, and the model with the lowest Akaike information criterion (AIC) was selected.

Table S14. List of the GenBank accession numbers for RSV consensus sequences used in phylogenetic analyses.

| **RSV-A (N = 207)** | | | **RSV-B (N = 176)** | | |
|---|---|---|---|---|---|
| LR699315 | MZ515752 | MZ515957 | LR699726 | MZ515711 | MZ515889 |
| LR699734 | MZ515766 | MZ515958 | LR699735 | MZ515712 | MZ515895 |
| LR699736 | MZ515773 | MZ515959 | LR699738 | MZ515714 | MZ515903 |
| LR699737 | MZ515777 | MZ515960 | LR699739 | MZ515715 | MZ515904 |
| MZ515551 | MZ515780 | MZ515961 | LR699740 | MZ515719 | MZ515918 |
| MZ515555 | MZ515782 | MZ515962 | LR699741 | MZ515724 | MZ515924 |
| MZ515556 | MZ515784 | MZ515963 | LR699742 | MZ515725 | MZ515925 |
| MZ515559 | MZ515788 | MZ515966 | LR699743 | MZ515727 | MZ515926 |
| MZ515566 | MZ515789 | MZ515967 | LR699744 | MZ515728 | MZ515930 |
| MZ515567 | MZ515790 | MZ515968 | MZ515553 | MZ515732 | MZ515938 |
| MZ515568 | MZ515800 | MZ515969 | MZ515554 | MZ515733 | MZ515944 |
| MZ515569 | MZ515801 | MZ515971 | MZ515557 | MZ515738 | MZ515946 |
| MZ515570 | MZ515802 | MZ515983 | MZ515558 | MZ515743 | MZ515947 |
| MZ515571 | MZ515803 | MZ515984 | MZ515560 | MZ515745 | MZ515950 |
| MZ515572 | MZ515821 | MZ515985 | MZ515562 | MZ515747 | MZ515953 |
| MZ515573 | MZ515825 | MZ515987 | MZ515563 | MZ515748 | MZ515955 |
| MZ515575 | MZ515828 | MZ515992 | MZ515565 | MZ515751 | MZ515970 |
| MZ515577 | MZ515833 | MZ515993 | MZ515574 | MZ515756 | MZ515972 |
| MZ515582 | MZ515834 | MZ515994 | MZ515578 | MZ515761 | MZ515975 |
| MZ515583 | MZ515835 | MZ516000 | MZ515581 | MZ515762 | MZ515991 |
| MZ515592 | MZ515840 | MZ516002 | MZ515584 | MZ515765 | MZ515997 |
| MZ515597 | MZ515841 | MZ516005 | MZ515586 | MZ515769 | MZ516003 |
| MZ515604 | MZ515842 | MZ516008 | MZ515590 | MZ515770 | MZ516006 |
| MZ515606 | MZ515848 | MZ516011 | MZ515591 | MZ515771 | MZ516016 |
| MZ515609 | MZ515850 | MZ516012 | MZ515595 | MZ515775 | MZ516020 |
| MZ515614 | MZ515851 | MZ516014 | MZ515598 | MZ515776 | MZ516021 |
| MZ515616 | MZ515852 | MZ516015 | MZ515599 | MZ515779 | MZ516022 |
| MZ515617 | MZ515854 | MZ516017 | MZ515603 | MZ515785 | MZ516025 |
| MZ515618 | MZ515859 | MZ516024 | MZ515605 | MZ515786 | MZ516030 |
| MZ515619 | MZ515860 | MZ516026 | MZ515607 | MZ515793 | MZ516041 |

| **RSV-A (N = 207)** | | | **RSV-B (N = 176)** | | |
|---|---|---|---|---|---|
| MZ515620 | MZ515861 | MZ516027 | MZ515608 | MZ515794 | MZ516042 |
| MZ515628 | MZ515862 | MZ516028 | MZ515610 | MZ515807 | MZ516049 |
| MZ515629 | MZ515866 | MZ516029 | MZ515612 | MZ515809 | MZ516051 |
| MZ515631 | MZ515875 | MZ516031 | MZ515613 | MZ515812 | MZ516054 |
| MZ515632 | MZ515876 | MZ516033 | MZ515615 | MZ515813 | MZ516056 |
| MZ515634 | MZ515877 | MZ516038 | MZ515625 | MZ515817 | MZ516060 |
| MZ515640 | MZ515878 | MZ516039 | MZ515627 | MZ515818 | MZ516061 |
| MZ515643 | MZ515881 | MZ516040 | MZ515636 | MZ515820 | MZ516062 |
| MZ515645 | MZ515882 | MZ516043 | MZ515637 | MZ515823 | MZ516065 |
| MZ515647 | MZ515884 | MZ516044 | MZ515638 | MZ515824 | MZ516081 |
| MZ515649 | MZ515887 | MZ516047 | MZ515639 | MZ515827 | MZ516095 |
| MZ515650 | MZ515896 | MZ516048 | MZ515653 | MZ515829 | MZ516098 |
| MZ515651 | MZ515898 | MZ516052 | MZ515656 | MZ515830 | MZ516102 |
| MZ515652 | MZ515901 | MZ516053 | MZ515658 | MZ515831 | MZ516104 |
| MZ515655 | MZ515902 | MZ516057 | MZ515660 | MZ515832 | MZ516105 |
| MZ515679 | MZ515905 | MZ516058 | MZ515663 | MZ515836 | MZ516107 |
| MZ515680 | MZ515907 | MZ516068 | MZ515665 | MZ515837 | MZ516109 |
| MZ515681 | MZ515908 | MZ516072 | MZ515667 | MZ515839 | MZ516113 |
| MZ515682 | MZ515909 | MZ516073 | MZ515669 | MZ515843 | MZ516114 |
| MZ515685 | MZ515910 | MZ516075 | MZ515671 | MZ515845 | MZ516119 |
| MZ515686 | MZ515911 | MZ516076 | MZ515672 | MZ515846 | MZ516122 |
| MZ515688 | MZ515912 | MZ516077 | MZ515674 | MZ515849 | MZ516123 |
| MZ515689 | MZ515913 | MZ516078 | MZ515691 | MZ515855 | MZ516135 |
| MZ515692 | MZ515921 | MZ516080 | MZ515698 | MZ515863 | MZ516136 |
| MZ515696 | MZ515922 | MZ516088 | MZ515699 | MZ515865 | MZ516139 |
| MZ515701 | MZ515923 | MZ516090 | MZ515704 | MZ515869 | MZ516140 |
| MZ515703 | MZ515928 | MZ516092 | MZ515707 | MZ515871 | MZ516141 |
| MZ515706 | MZ515929 | MZ516099 | MZ515708 | MZ515872 | MZ516143 |
| MZ515717 | MZ515931 | MZ516100 | MZ515710 | MZ515883 | OP963385 |
| MZ515718 | MZ515933 | MZ516103 | | | |
| MZ515720 | MZ515939 | MZ516108 | | | |
| MZ515722 | MZ515940 | MZ516110 | | | |

| RSV-A (N = 207) | | | RSV-B (N = 176) |
|---|---|---|---|
| MZ515723 | MZ515941 | MZ516112 | |
| MZ515731 | MZ515942 | MZ516117 | |
| MZ515734 | MZ515943 | MZ516120 | |
| MZ515740 | MZ515945 | MZ516129 | |
| MZ515741 | MZ515949 | MZ516132 | |
| MZ515744 | MZ515951 | MZ516134 | |
| MZ515749 | MZ515956 | MZ516137 | |

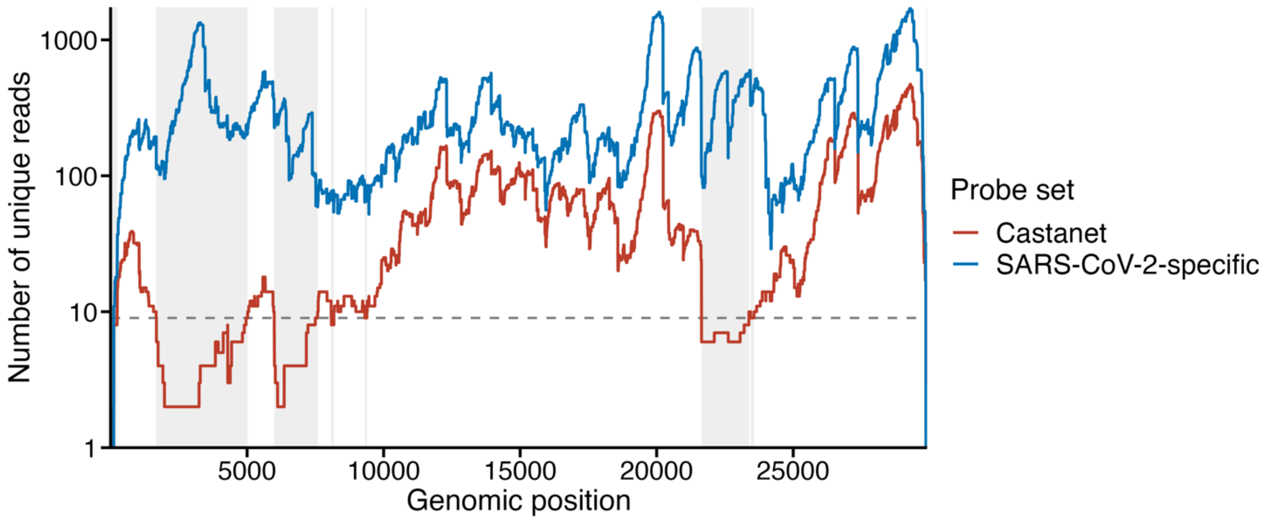

Figure S1. Coverage plot for SARS-CoV-2. In a previous study[1], a sample collected from a patient infected with severe acute respiratory syndrome coronavirus 2 (SARS-CoV-2) was sequenced using targeted metagenomic sequencing with the Castanet probe set (red) and a SARS-CoV-2-specific probe set (blue). The SARS-CoV-2 genome was reconstructed as previously described[1]. Genomic regions with coverage of <10 Castanet-captured unique reads were shaded in grey (22.7 kb). The dashed grey line represents coverage of nine unique reads.

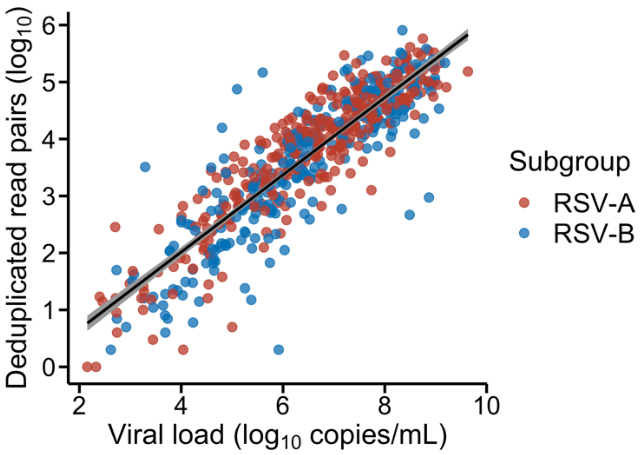

Figure S2. The correlation between the number of unique RSV reads and RSV viral load. Viral load was measured using reverse transcription quantitative PCR in 621 samples. On average, for every $\log_{10}$ increase in read count, viral load increases by 1.2 $\log_{10}$. Samples with both RSV subgroups detected were excluded from this figure (N = 9).

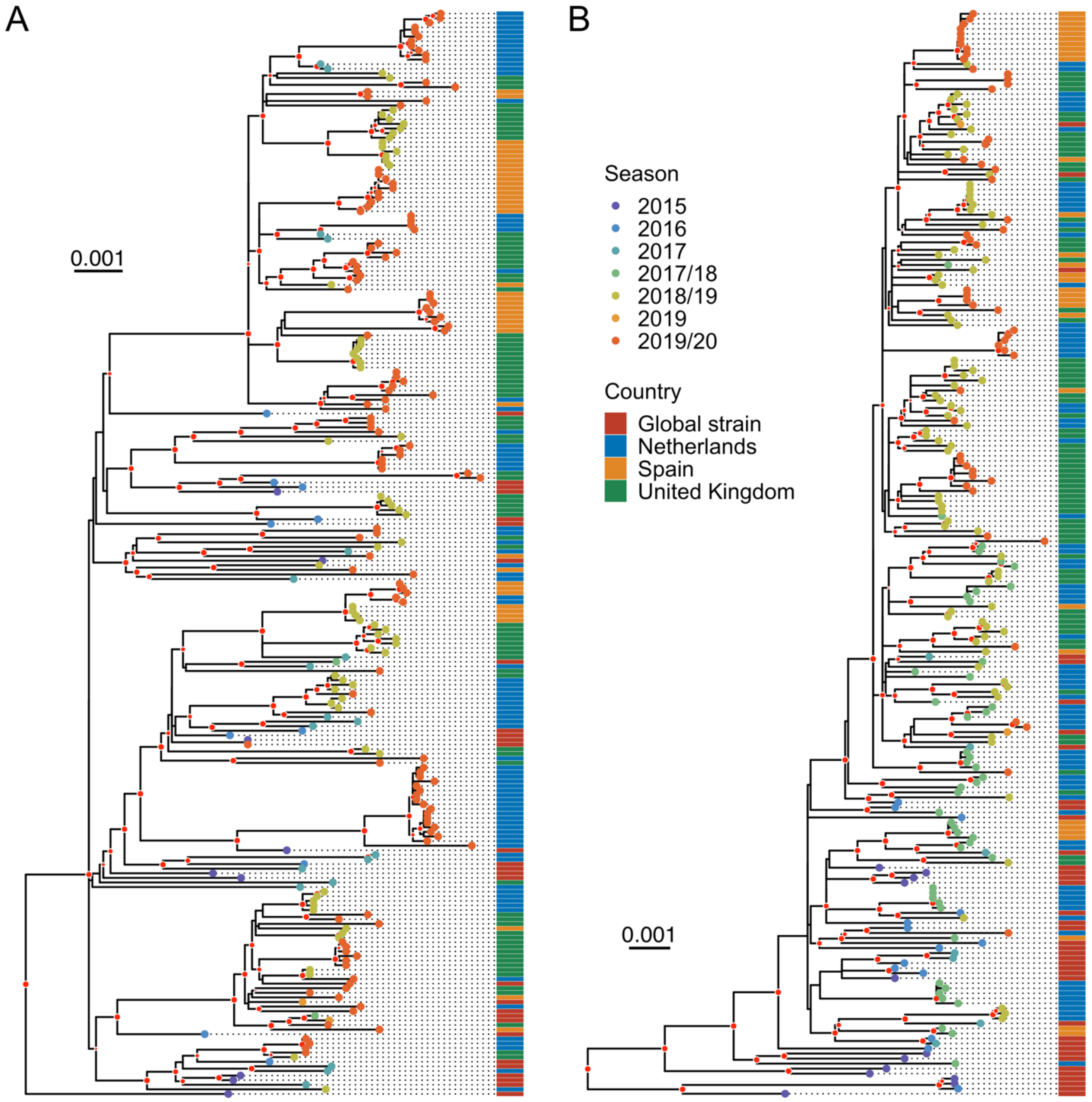

Figure S3. Maximum-likelihood phylogenies of strains from the present study and strains collected from all over the world between 2015 and 2019, downloaded from GenBank. A. RSV-A phylogeny. All RSV-A strains from the present study and GenBank were genotype ON1, except for the one at the very bottom, which is genotype GA2, downloaded from GenBank (accession number MH181907). B. RSV-B phylogeny. All RSV-B strains from the present study and GenBank were genotype BA. On average, ON1 strains had greater patristic distances than BA ones (mean ± standard deviation, 0.0100 ± 0.0028 vs. 0.0063 ± 0.0030; Mann–Whitney U test, $P < 2.2\times10^{-16}$), whereas BA strains showed stronger temporal clustering than ON1 ones. The phylogenies were inferred using RAxML with the general time reversible nucleotide substitution model and gamma-distributed rate heterogeneity among sites. The sampling season and country of each strain are illustrated by tip and bar colours. Strains labelled as Netherlands, Spain, and United Kingdom were generated from this study, and the remainder were downloaded from GenBank. Red dots show well-supported nodes with a bootstrap value over 70% and are sized in proportion to bootstrap values. The trees were rooted to the oldest strains sampled during 1962–1977—GenBank accession numbers KU316149 and KU316166 for RSV-A, and KU316116 and MG813995 for RSV-B. The oldest strains are not shown here. The scale bars represent the number of nucleotide substitutions per site.

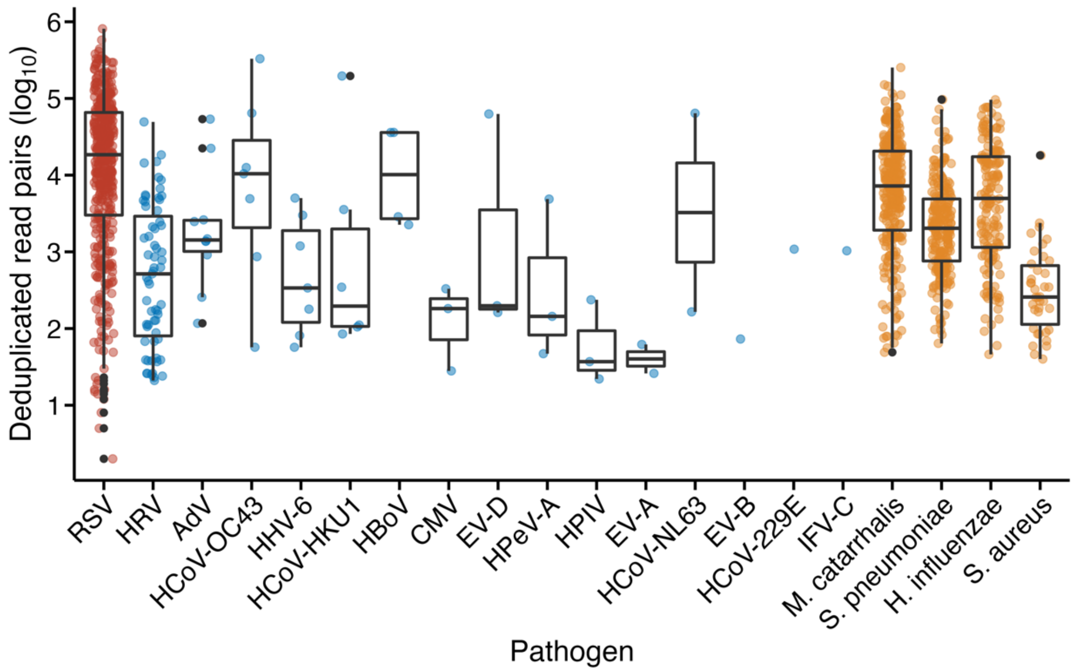

Figure S4. Number of deduplicated read pairs mapped to each pathogen detected in this study. The centre line of each box denotes the median; box limits, the first and third quartiles; whiskers, the highest and lowest values within 1.5 times the interquartile range from the box limits; and outlying points, outliers. The species from left to right are: respiratory syncytial virus (RSV), human rhinovirus (HRV), adenovirus (AdV), human coronavirus OC43 (HCoV-OC43), human herpesvirus 6 (HHV-6), human coronavirus HKU1 (HCoV-HKU1), human bocavirus (HBoV), cytomegalovirus (CMV), enterovirus D (EV-D), human parechovirus A (HPeV-A), human parainfluenza virus (HPIV), enterovirus A (EV-A), human coronavirus NL63 (HCoV-NL63), enterovirus B (EV-B), human coronavirus 229E (HCoV-229E), influenza C virus (IFV-C), *Moraxella catarrhalis*, *Streptococcus pneumoniae*, *Haemophilus influenzae*, and *Staphylococcus aureus*.

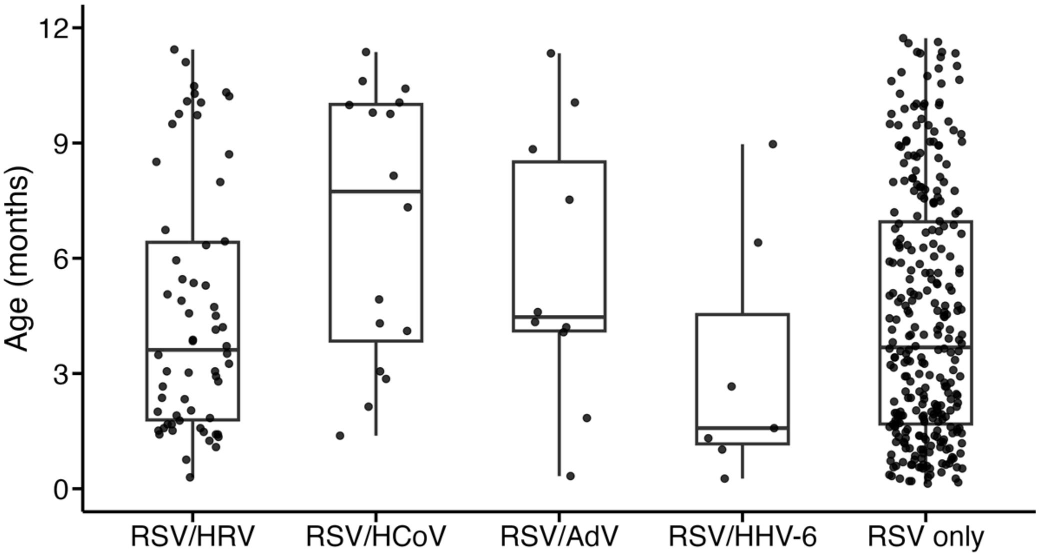

Figure S5. Age distribution among RSV-infected infants with and without co-detection of any other virus. The age distribution between these groups was significantly different (Kruskal–Wallis test, P = 0.030). Infants with RSV/human coronaviruses (HCoV) co-detection were significantly older than those with RSV/human herpesvirus 6 (HHV-6) co-detection (Dunn's test with the Benjamini–Hochberg method, adjusted P = 0.044). The median (interquartile range) age of infants with RSV/human rhinovirus (HRV), RSV/HCoV, RSV/adenoviruses (AdV), RSV/HHV-6, and RSV only was 3.6 (1.8–6.4), 7.7 (3.8–10.0), 4.5 (4.1–8.5), 1.6 (1.2–4.5), and 3.7 (1.7–6.9) months, respectively. The centre line of each box denotes the median; box limits, the first and third quartiles; and whiskers, the highest and lowest values within 1.5 times the interquartile range from the box limits.

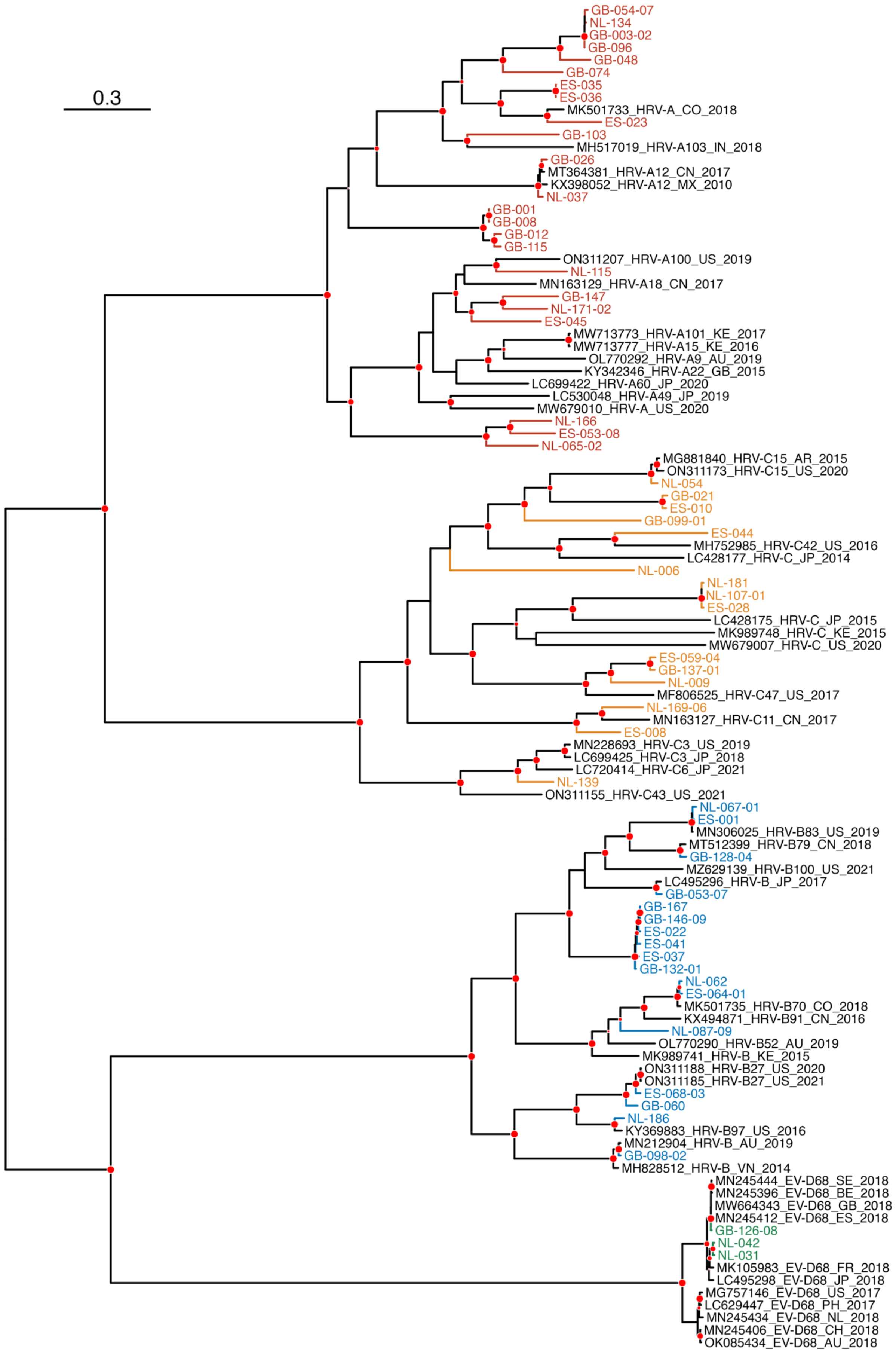
0.3
GB-054-07
NL-134
GB-003-02
GB-096
GB-048
GB-074
ES-035
ES-036
MK501733_HRV-A_CO_2018
ES-023
GB-103
MH517019_HRV-A103_IN_2018
GB-026
MT364381_HRV-A12_CN_2017
KX398052_HRV-A12_MX_2010
NL-037
GB-001
GB-008
GB-012
GB-115
ON311207_HRV-A100_US_2019
NL-115
MN163129_HRV-A18_CN_2017
GB-147
NL-171-02
ES-045
MW713773_HRV-A101_KE_2017
MW713777_HRV-A15_KE_2016
OL770292_HRV-A9_AU_2019
KY342346_HRV-A22_GB_2015
LC699422_HRV-A60_JP_2020
LC530048_HRV-A49_JP_2019
MW679010_HRV-A_US_2020
NL-166
ES-053-08
NL-065-02
MG881840_HRV-C15_AR_2015
ON311173_HRV-C15_US_2020
NL-054
GB-021
ES-010
GB-099-01
ES-044
MH752985_HRV-C42_US_2016
LC428177_HRV-C_JP_2014
NL-006
NL-181
NL-107-01
ES-028
LC428175_HRV-C_JP_2015
MK989748_HRV-C_KE_2015
MW679007_HRV-C_US_2020
ES-059-04
GB-137-01
NL-009
MF806525_HRV-C47_US_2017
NL-169-06
MN163127_HRV-C11_CN_2017
ES-008
MN228693_HRV-C3_US_2019
LC699425_HRV-C3_JP_2018
LC720414_HRV-C6_JP_2021
NL-139
ON311155_HRV-C43_US_2021
NL-067-01
ES-001
MN306025_HRV-B83_US_2019
MT512399_HRV-B79_CN_2018
GB-128-04
MZ629139_HRV-B100_US_2021
LC495296_HRV-B_JP_2017
GB-053-07
GB-167
GB-146-09
ES-022
ES-041
ES-037
GB-132-01
NL-062
ES-064-01
MK501735_HRV-B70_CO_2018
KX494871_HRV-B91_CN_2016
NL-087-09
OL770290_HRV-B52_AU_2019
MK989741_HRV-B_KE_2015
ON311188_HRV-B27_US_2020
ON311185_HRV-B27_US_2021
ES-068-03
GB-060
NL-186
KY369883_HRV-B97_US_2016
MN212904_HRV-B_AU_2019
GB-098-02
MH828512_HRV-B_VN_2014
MN245444_EV-D68_SE_2018
MN245396_EV-D68_BE_2018
MW664343_EV-D68_GB_2018
MN245412_EV-D68_ES_2018
GB-126-08
NL-042
NL-031
MK105983_EV-D68_FR_2018
LC495298_EV-D68_JP_2018
MG757146_EV-D68_US_2017
LC629447_EV-D68_PH_2017
MN245434_EV-D68_NL_2018
MN245406_EV-D68_CH_2018
OK085434_EV-D68_AU_2018

Figure S6. Phylogenetic reconstruction of genus *Enterovirus* recovered from this study. Samples where less than 50% of the genome was recovered were removed from the reconstruction. For infants who had multiple samples collected, only the sample with the highest number of *Enterovirus* reads was included. The samples from this study were further supplemented with 50 complete or nearly complete *Enterovirus* genomes downloaded from GenBank, collected from across the globe between 2010 and 2021. RAxML was used to reconstruct the maximum-likelihood phylogeny with the general time reversible nucleotide substitution model and gamma-distributed rate heterogeneity among sites. The tree was midpoint rooted. Samples from this study are shown in different colours: *Rhinovirus A* (red), *Rhinovirus B* (blue), *Rhinovirus C* (tangerine), and *Enterovirus D* (green). All *Enterovirus D* strains are enterovirus D68. Samples were labelled as anonymised subject ID (2 letters followed by 3 digits) and order of serial samples if applicable. Strains downloaded from GenBank are shown in black, labelled as GenBank accession number, followed by type, sampling country, and sampling year. Red dots show well-supported nodes with a bootstrap value over 70% and are sized in proportion to bootstrap values. The scale bars represent the number of nucleotide substitutions per site. AR denotes Argentina; AU, Australia; BE, Belgium; CH, Switzerland; CN, China; CO, Colombia; ES, Spain; FR, France; GB, United Kingdom; IN, India; JP, Japan; KE, Kenya; MX, Mexico; NL, Netherlands; PH, Philippines; SE, Sweden; US, United States; VN, Viet Nam.

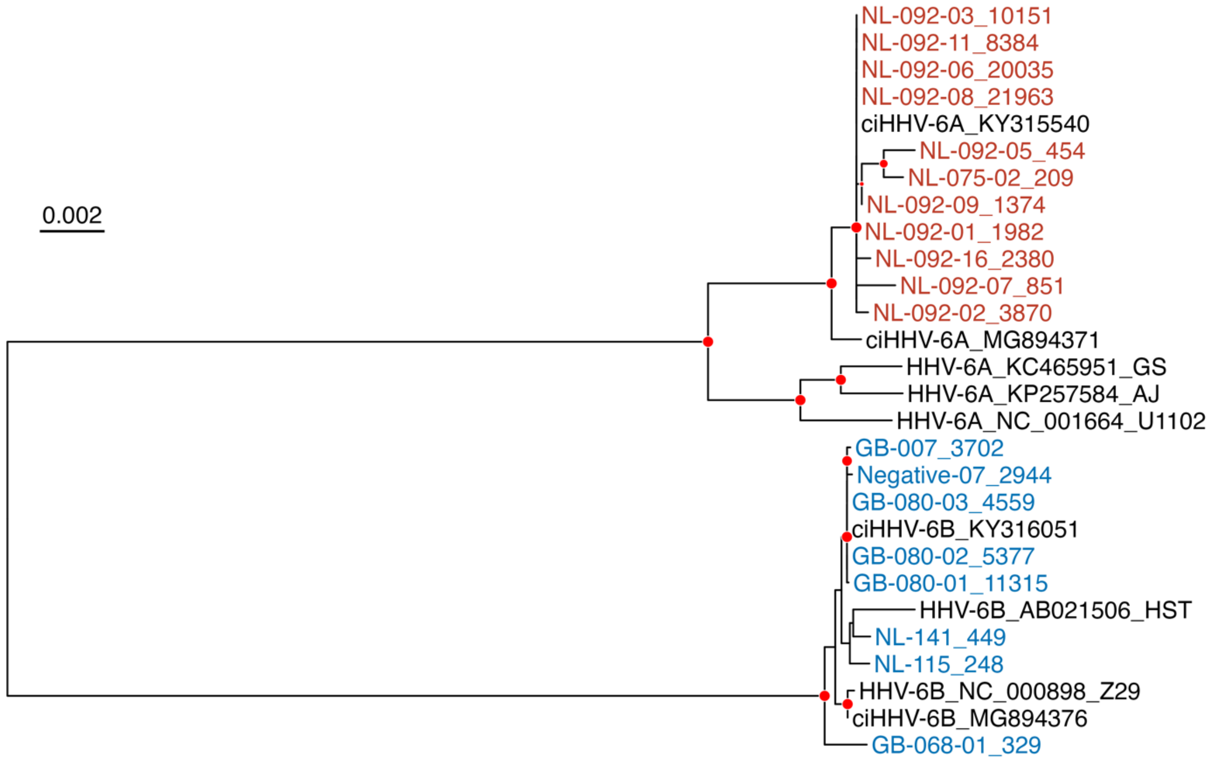

Figure S7. Phylogenetic reconstruction of human herpesvirus 6 (HHV-6) in samples from this study, contextualised with HHV-6 reference genomes. RAxML was used to reconstruct the maximum-likelihood phylogeny with the general time reversible nucleotide substitution model and gamma-distributed rate heterogeneity among sites. Coloured taxa are samples collected from this study, HHV-6A in red and HHV-6B in blue. They are labelled as anonymised patient ID (2 letters followed by 3 digits), order of serial samples if applicable, and number of HHV-6 reads, except for one taxon labelled as Negative-07_2944, which had 2,944 HHV-6 reads and was collected from a participant who had respiratory symptoms but tested negative for RSV. Taxa in black represent the reference genomes, labelled as HHV-6 type (ci denotes chromosomally integrated), GenBank accession number, and strain name if applicable. The tree was midpoint rooted. Red dots show well-supported nodes with a bootstrap value over 70% and are sized in proportion to bootstrap values. The scale bar represents the number of nucleotide substitutions per site.

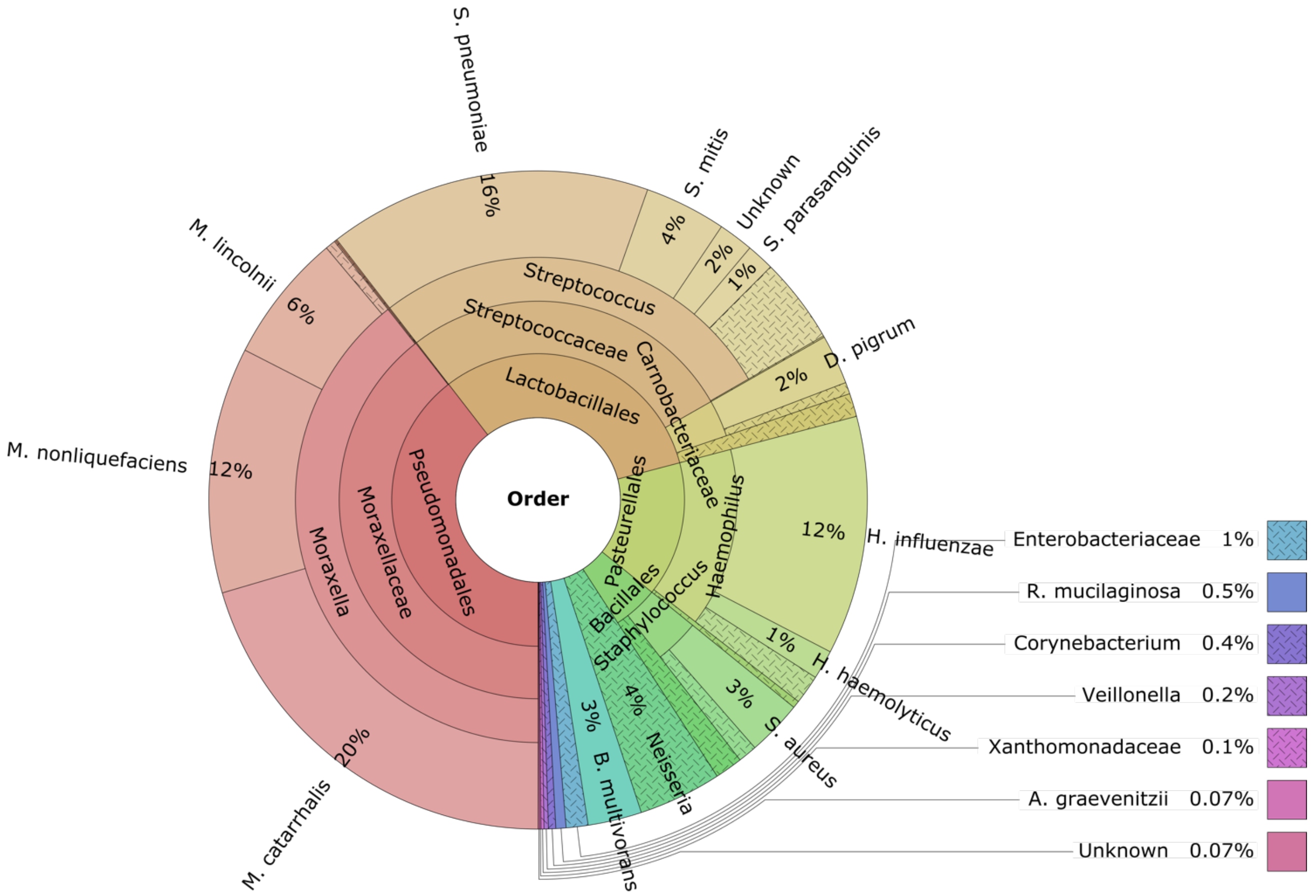

Figure S8. Composition of the bacteria identified in the nasopharynx of the RSV-infected infants. The percentage of a bacterium was calculated as the number of infants having this bacterium divided by the sum of the number of bacterial species found in each infant. Circles from inside to outside represent the order, family, genus, and species of the bacteria with some levels collapsed for better visualisation. Interactive visualisation can be accessed in the Supplementary html file.

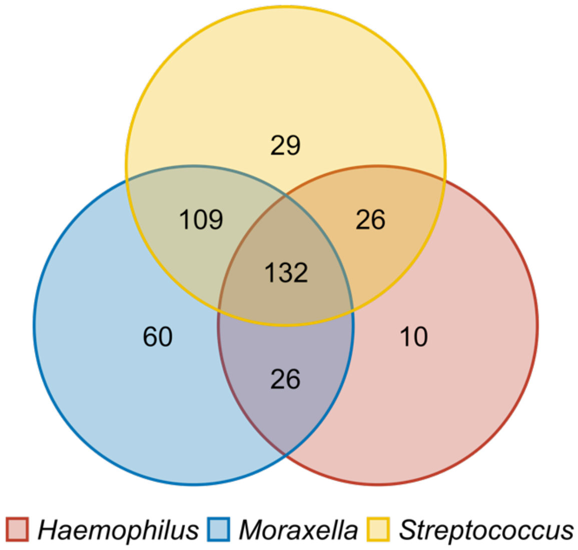

Figure S9. Venn diagram of the three main bacterial genera found in RSV-infected infants. The number in each area represents the number of infants. Forty-one infants did not have any of the three bacteria.

A

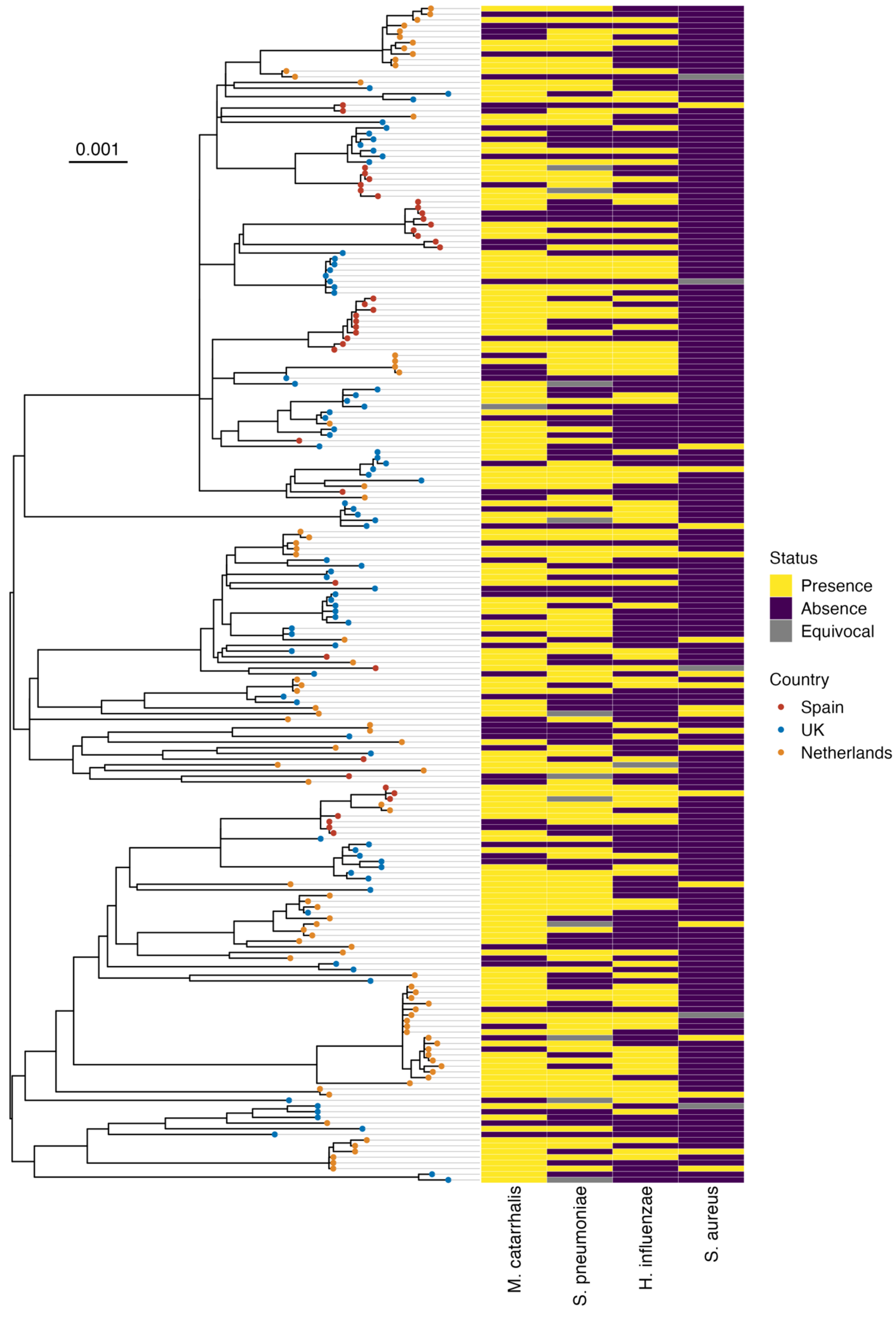
0.001
M. catarrhalis
S. pneumoniae
H. influenzae
S. aureus
Status
Presence
Absence
Equivocal
Country
Spain
UK
Netherlands

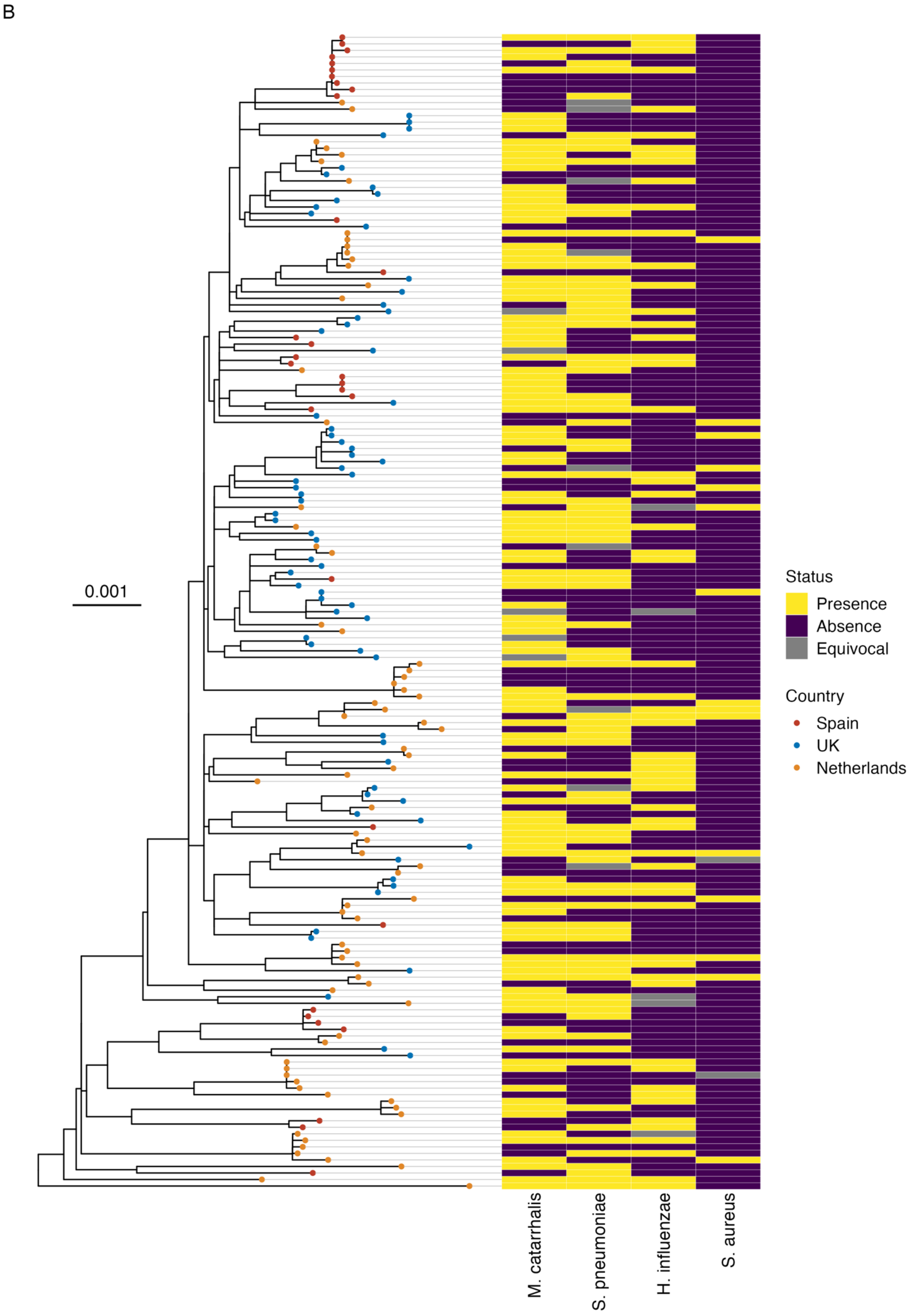
B
0.001
M. catarrhalis
S. pneumoniae
H. influenzae
S. aureus
Status
Presence
Absence
Equivocal
Country
Spain
UK
Netherlands

Figure S10. Distribution of commonly co-detected bacteria on the (A) RSV-A and (B) RSV-B maximum-likelihood phylogenies. The phylogenies were reconstructed from samples in which at least 70% of the RSV coding sequences were recovered, using RAxML with the general time reversible nucleotide substitution model and gamma-distributed rate heterogeneity among sites. For infants with multiple samples collected, only the sample with the highest coverage was included. The trees were midpoint rooted. The sampling country of each strain is illustrated by tip colour. The scale bars represent the number of nucleotide substitutions per site. *M. catarrhalis* denotes *Moraxella catarrhalis*; *S. pneumoniae*, *Streptococcus pneumoniae*; *H. influenzae*, *Haemophilus influenzae*; *S. aureus*, *Staphylococcus aureus*.

**References**

1. Lythgoe KA, Hall M, Ferretti L, et al. SARS-CoV-2 within-host diversity and transmission. Science 2021;372:eabg0821.
